# Mapping the aetiological foundations of the heart failure spectrum using human genetics

**DOI:** 10.1101/2023.10.01.23296379

**Authors:** Albert Henry, Xiaodong Mo, Chris Finan, Mark D. Chaffin, Doug Speed, Hanane Issa, Spiros Denaxas, James S. Ware, Sean L. Zheng, Anders Malarstig, Jasmine Gratton, Isabelle Bond, Carolina Roselli, David Miller, Sandesh Chopade, A. Floriaan Schmidt, Erik Abner, Lance Adams, Charlotte Andersson, Krishna G. Aragam, Johan Ärnlöv, Geraldine Asselin, Anna Axelsson Raja, Joshua D. Backman, Traci M. Bartz, Kiran J. Biddinger, Mary L. Biggs, Heather L. Bloom, Eric Boersma, Jeffrey Brandimarto, Michael R. Brown, Søren Brunak, Mie Topholm Bruun, Leonard Buckbinder, Henning Bundgaard, David J. Carey, Daniel I. Chasman, Xing Chen, James P. Cook, Tomasz Czuba, Simon de Denus, Abbas Dehghan, Graciela E. Delgado, Alexander S. Doney, Marcus Dörr, Joseph Dowsett, Samuel C. Dudley, Gunnar Engström, Christian Erikstrup, Tõnu Esko, Eric H. Farber-Eger, Stephan B. Felix, Sarah Finer, Ian Ford, Mohsen Ghanbari, Sahar Ghasemi, Jonas Ghouse, Vilmantas Giedraitis, Franco Giulianini, John S. Gottdiener, Stefan Gross, Daníel F. Guðbjartsson, Hongsheng Gui, Rebecca Gutmann, Sara Hägg, Christopher M. Haggerty, Åsa K. Hedman, Anna Helgadottir, Harry Hemingway, Hans Hillege, Craig L. Hyde, Bitten Aagaard Jensen, J. Wouter Jukema, Isabella Kardys, Ravi Karra, Maryam Kavousi, Jorge R. Kizer, Marcus E. Kleber, Lars Køber, Andrea Koekemoer, Karoline Kuchenbaecker, Yi-Pin Lai, David Lanfear, Claudia Langenberg, Honghuang Lin, Lars Lind, Cecilia M. Lindgren, Peter P. Liu, Barry London, Brandon D. Lowery, Jian’an Luan, Steven A. Lubitz, Patrik Magnusson, Kenneth B. Margulies, Nicholas A. Marston, Hilary Martin, Winfried März, Olle Melander, Ify R. Mordi, Michael P. Morley, Andrew P. Morris, Alanna C. Morrison, Lori Morton, Michael W. Nagle, Christopher P. Nelson, Alexander Niessner, Teemu Niiranen, Raymond Noordam, Christoph Nowak, Michelle L. O’Donoghue, Sisse Rye Ostrowski, Anjali T. Owens, Colin N. A. Palmer, Guillaume Paré, Ole Birger Pedersen, Markus Perola, Marie Pigeyre, Bruce M. Psaty, Kenneth M. Rice, Paul M. Ridker, Simon P. R. Romaine, Jerome I. Rotter, Christian T. Ruff, Mark S. Sabatine, Neneh Sallah, Veikko Salomaa, Naveed Sattar, Alaa A. Shalaby, Akshay Shekhar, Diane T. Smelser, Nicholas L. Smith, Erik Sørensen, Sundararajan Srinivasan, Kari Stefansson, Garðar Sveinbjörnsson, Per Svensson, Mari-Liis Tammesoo, Jean-Claude Tardif, Maris Teder-Laving, Alexander Teumer, Guðmundur Thorgeirsson, Unnur Thorsteinsdottir, Christian Torp-Pedersen, Vinicius Tragante, Stella Trompet, Andre G. Uitterlinden, Henrik Ullum, Pim van der Harst, David van Heel, Jessica van Setten, Marion van Vugt, Abirami Veluchamy, Monique Verschuuren, Niek Verweij, Christoffer Rasmus Vissing, Uwe Völker, Adriaan A. Voors, Lars Wallentin, Yunzhang Wang, Peter E. Weeke, Kerri L. Wiggins, L. Keoki Williams, Yifan Yang, Bing Yu, Faiez Zannad, Chaoqun Zheng, Genes & Health Research Team, DBDS Genomic Consortium, Folkert W. Asselbergs, Thomas P. Cappola, Marie-Pierre Dubé, Michael E. Dunn, Chim C. Lang, Nilesh J. Samani, Svati Shah, Ramachandran S. Vasan, J. Gustav Smith, Hilma Holm, Sonia Shah, Patrick T. Ellinor, Aroon D. Hingorani, Quinn Wells, R. Thomas Lumbers, HERMES Consortium

## Abstract

Heart failure (HF), a syndrome of symptomatic fluid overload due to cardiac dysfunction, is the most rapidly growing cardiovascular disorder. Despite recent advances, mortality and morbidity remain high and treatment innovation is challenged by limited understanding of aetiology in relation to disease subtypes. Here we harness the de-confounding properties of genetic variation to map causal biology underlying the HF phenotypic spectrum, to inform the development of more effective treatments. We report a genetic association analysis in 1.9 million ancestrally diverse individuals, including 153,174 cases of HF; 44,012 of non-ischaemic HF; 5,406 cases of non-ischaemic HF with reduced ejection fraction (HFrEF); and 3,841 cases of non-ischaemic HF with preserved ejection fraction (HFpEF). We identify 66 genetic susceptibility loci across HF subtypes, 37 of which have not previously been reported. We map the aetiologic contribution of risk factor traits and diseases as well as newly identified effector genes for HF, demonstrating differential risk factor effects on disease subtypes. Our findings highlight the importance of extra-cardiac tissues in HF, particularly the kidney and the vasculature in HFpEF. Pathways of cellular senescence and proteostasis are notably uncovered, including *IGFBP7* as an effector gene for HFpEF. Using population approaches causally anchored in human genetics, we provide fundamental new insights into the aetiology of heart failure subtypes that may inform new approaches to prevention and treatment.

## Main

Genome-wide association studies (GWAS) of heart failure (HF) and cardiac traits have begun to yield important translational insights, however, existing studies are limited by phenotypic heterogeneity and a lack of stratification by major aetiologies^1–3^. To address these limitations, we performed a GWAS meta-analysis of 153,174 HF cases in 1.9 million ancestrally diverse individuals, including analysis of 4 HF subtypes defined by aetiology and left ventricular ejection fraction (**Supplementary Figure 1**). Multimodal definitions were used to harmonise the ascertainment of heart failure (hereafter referred to as HF), non-ischaemic HF (ni-HF), and non-ischaemic HF stratified by left ventricular ejection fraction (LVEF) below or above 50% (ni-HFrEF and ni-HFpEF respectively), together with corresponding control populations. We characterise the common genetic architecture of HF subtypes and use this information to identify the key effector tissues, pathways and genes underlying aetiology. We map the genetic pleiotropy of HF genomic risk loci to identify aetiological phenotype clusters and use Mendelian randomisation to estimate subtype-specific effects of cardiometabolic traits and diseases. These findings provide new insights into the aetiology of HF subtypes and provide a basis for the development of new therapeutic approaches.

## Results

### Multi-ancestry genetic association analysis highlights novel heart failure loci

We performed a meta-analysis of case-control GWAS across 42 studies to investigate the association of up to 10,199,961 common genetic variants (minor allele frequency (MAF) >1%) with risk of four HF phenotypes. The study population comprised 1,946,349 individuals representing five major ancestry groups: European (86.6%), African (0.8%), South Asian (1.5%), East Asian (10.9%), and Hispanic (0.1%). 153,174 HF cases were identified, including 44,012 cases of non-ischaemic HF; 5,406 cases of non-ischaemic HFrEF; and 3,841 cases of non-ischaemic HFpEF (**Supplementary Figure 2**). We identified 59 conditionally independent (sentinel) genetic variants at 56 non-overlapping genomic loci (distance > 500 kilobase pairs) associated with HF at a genome-wide significance (*P* < 5 x 10^-8^) (**Figure 1, Supplementary Table 1**). Sentinel variants showed consistent allelic effects across ancestry groups, except one variant at the *LPA* locus (*P*_*het-ancestry*_ < 0.05 / 59) (**Supplementary Figure 3, Supplementary Table 2**). GWAS of ni-HF, ni-HFrEF, and ni-HFpEF subtypes highlighted 10 additional sentinel variants that were not identified in the HF GWAS. In total, 66 independent genomic risk loci were identified across HF phenotypes^1,4,5^ among which 37 have not previously been reported in GWAS of HF^1–5^. Of the 66 loci, 46 (70%) were associated with at least one of the non-ischaemic HF phenotypes at *P* < 0.05 / 66. These loci were then classified as non-ischaemic HF loci, and the remaining 20 (30%) HF loci were classified as secondary HF (s-HF) loci. Amongst the non-ischaemic loci, 17 were associated with ni-HFrEF and 3 were associated with ni-HFpEF at *P* < 0.05 / 66. In addition, we replicated 76 / 87 (87%) previously reported HF sentinel variants that were available in our study (*P* < 0.05 / 87) (**Supplementary Figure 4, Supplementary Table 3)**.

**Figure 1.**
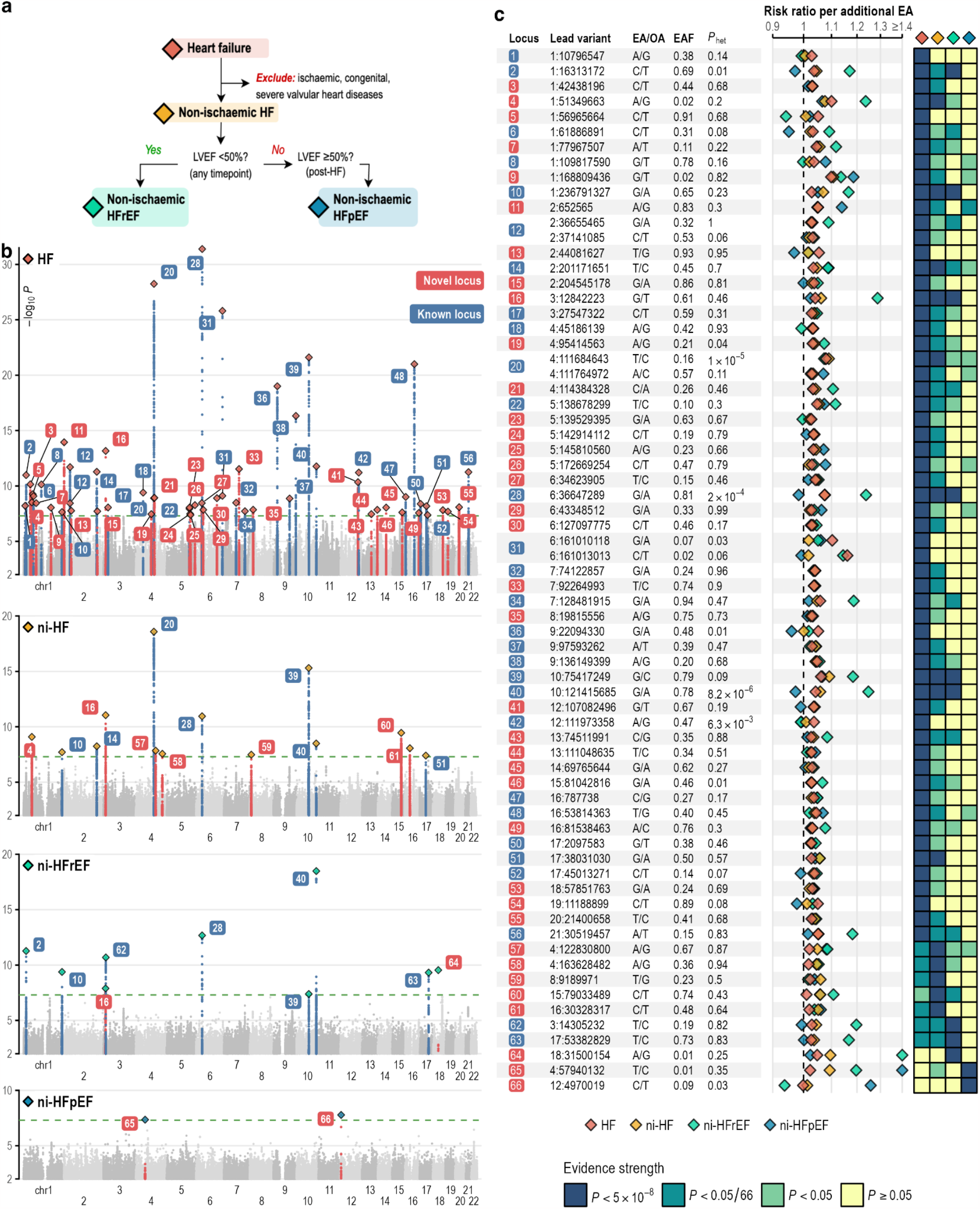
**a**. Phenotyping schema **b**. Manhattan plots of 4 HF subtypes. **c**. Summary of conditionally independent lead variants across HF phenotypes. Lead variants are denoted using chromosome and base pair position according to GRCh37 assembly. RR = risk ratio, EA = effect allele, aligned to risk-increasing allele in HF phenotype with lowest *P* value for association; OA = other (non-effect) allele; EAF = effect allele frequency, *P*_*het*_ = *P* value for effect estimate heterogeneity across studies. EAF and *P*_*het*_ estimates were taken from the first HF phenotype with genome-wide significant lead variant.

### Genetic architecture and heritability heart failure subtypes

The genetic architecture of HF was found to be highly polygenic, evidenced by an elevated genomic inflation factor (*λ*_*GC*_ = 1.22) in the absence of population stratification (linkage disequilibrium score [LDSC] regression intercept = 1.01) **(Supplementary Figures 5-6)**, and there was an exponential relationship between allele frequency and effect size for associated variants (**Supplementary Figure 7**). The estimated proportion of variance in disease liability explained by common genetic variants, i.e. SNP-based heritability (*h*^2^_g_), was 5.4 ± 0.2% for HF; 6.1 ± 0.5% for non-ischaemic HF; 11.8 ± 2.6% for non-ischaemic HFrEF, and 1.8 ± 1.3% for non-ischaemic HFpEF (**Supplementary Figure 8**)^6^. All HF phenotype pairs demonstrated positive genetic correlations, with estimates ranging from 0.42 (standard error, SE = 0.18) between ni-HFrEF and ni-HFpEF to 0.93 (SE = 0.15) between ni-HF and ni-HFpEF (**Supplementary Figure 9**). The genetic correlation between ni-HFrEF and both HF (0.66, SE = 0.06) and ni-HF (0.74, SE = 0.06) was lower than for ni-HFpEF (0.85, SE = 0.14 for HF and 0.93, SE = 0.15 for ni-HF), suggesting heritable components that are specific to ni-HFrEF.

To explore the potential utility of SNP-based heritability for prediction, we derived a polygenic score (PGS_HF_) from the HF GWAS excluding UK Biobank (UKB) samples and evaluated the association with HF in UKB. Among 346,667 UKB European participants (13,824 HF cases), the PGS_HF_ was associated with HF (OR per PGS SD 1.37 [95% CI 1.27 to 1.46], P<2x10^-16^) after adjusting for sex, age and first 10 ten genetic principal components (PC)s. Individuals in the top PGS_HF_ decile had a 1.76-fold higher odds of developing heart failure compared to those in the fifth decile (OR = 1.76, 95% CI = 1.64 to 1.89) and 2.89-fold compared to those in the bottom decile (OR = 2.89, 95% CI = 2.66 to 3.14) (**Supplementary Figure 10**).

### Prioritisation of effector genes for heart failure

To identify HF effector genes, we characterised the functional properties of variants and genes within each identified GWAS locus using a range of orthogonal approaches. Functionally-informed fine-mapping^7^ identified 70 credible sets containing 547 likely causal variants (cumulative posterior inclusion probability > 0.95) at 47/66 HF loci. These credible sets included 11 single nucleotide variants with predicted deleteriousness ranked amongst the top 1% in the reference human genome (CADD Phred score >20)^8^, including exonic variants in established dilated cardiomyopathy (DCM) genes such as *FLNC, BAG3* and *HSPB7*^*9–11*^ (**Supplementary Figure 11, Supplementary Table 4**).

We then prioritised putative causal genes among 758 protein-coding genes overlapping HF loci by triangulating evidence from three predictors of gene relevance: 1) variant-to-gene (V2G) score derived from functional properties of fine-mapped and sentinel variants^12^, 2) polygenic priority score (PoPS) calculated from enrichment of gene features^13^, and 3) predicted gene expression levels across tissues derived from multi-tissue transcriptome-wide association study (TWAS)^14^. 142 genes were ranked by at least one predictor (labelled as *candidate genes*), of which 71 were further prioritised based on: ranking highest on a combined predictor-score; highest ranking with at least two predictors; colocalisation with gene transcript expression in a relevant tissue^15^; or association with a phenotypically relevant Mendelian disorder (labelled as *prioritised genes*)^16^ (**Figure 2a-c, Supplementary Table 5**).

**Figure 2.**
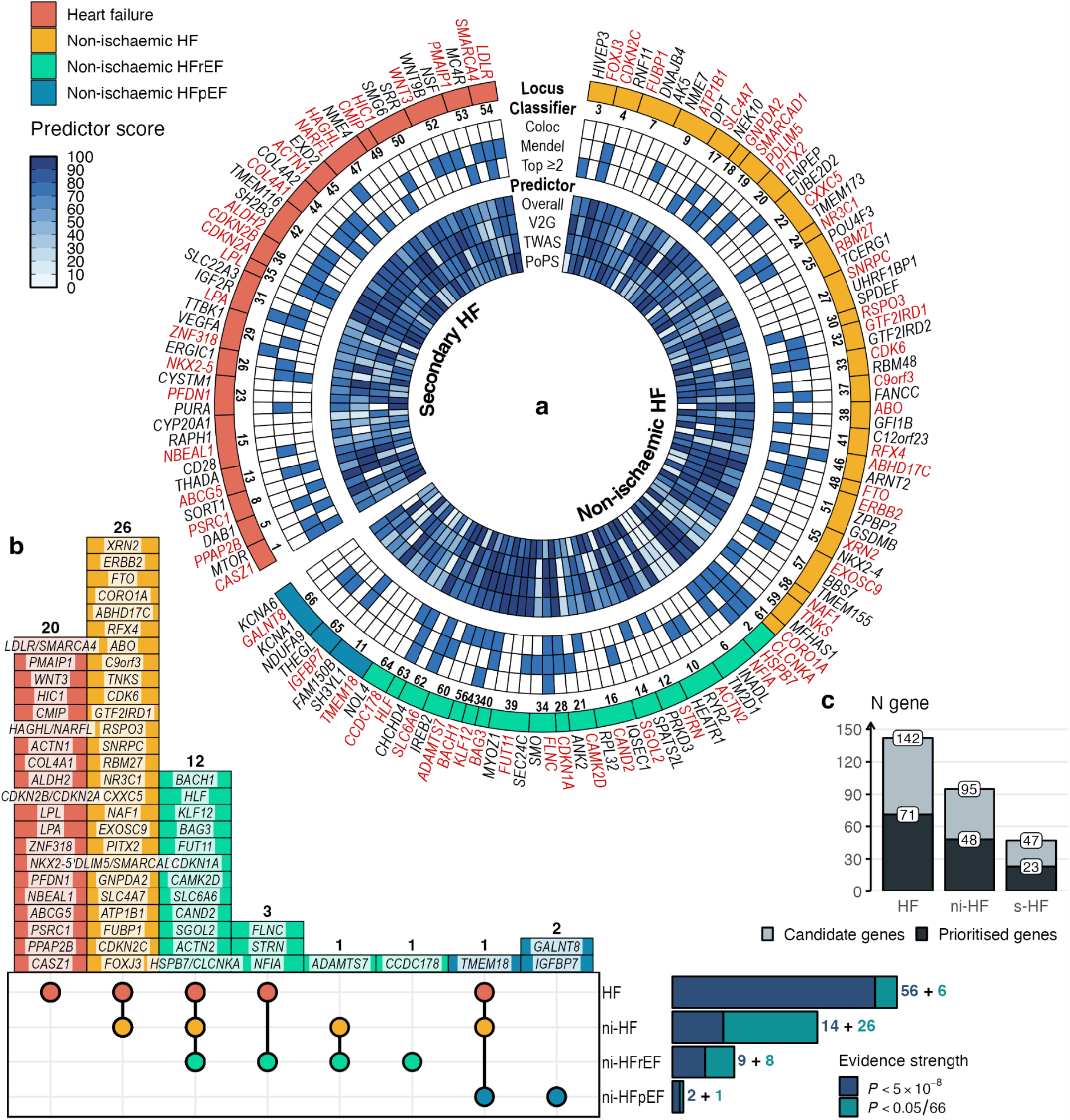
**a**. Sunburst chart of Candidate genes (black label) and Prioritised genes (red label) for HF identified through gene prioritisation strategy across 66 loci showing heatmap of predictor score and boolean classifier score (blue tile indicates a ‘True’ value). **b**. Upset plot showing number of loci categorised by phenotypic associations across HF subtypes, colour coded by the most specific association at P < 0.05 / 66 and labelled with Prioritised genes. **c**. Number of Candidate and Prioritised genes across HF phenotype gene set.

The prioritised genes at secondary HF loci included a high proportion with established evidence linking them to atherosclerotic diseases, for example via regulatory effects on cholesterol and lipoproteins metabolism *LDLR*^17^, *LPL*^18^, *ABCG5*^19,20^, *LPA*^21^, and *CDKN2A/CDKN2B*^*22*^ (commonly known as 9p21.3 locus). Prioritised genes at the non-ischaemic HF loci included established cardiomyopathy genes *BAG3, FLNC, ACTN2*, and *HSPB7*, ^9–11,23^, *CAMKD2* (linked to cardiac hypertrophy^24^), *STRN* (linked to canine DCM^25^), as well as *PITX2, KLF12*, and *ATP1B1* which have been associated with cardiac arrhythmia^26–28^. Other notable findings include *CAND2*, a muscle-specific gene of which upregulation has been linked to pathological cardiac remodelling^29^ and *NKX2-5*, a core cardiac transcription factor associated with congenital heart disease and cardiomyopathy^30–32^.

To facilitate genomic appraisal at each identified locus, we constructed an online dashboard visualisation containing regional genetic association, gene prioritisation, cross-trait association, and study-level estimate (**Supplementary Materials**).

### Biological pathways associated with heart failure subphenotypes

To identify convergent molecular disease mechanisms, we performed a gene set (candidate and prioritised) enrichment analysis. We identified 91 enriched biological pathways and cellular phenotypes for HF and 11 for non-ischaemic HF (adjusted *P* < 0.05) (**Figure 3a, Supplementary Table 6**). Several pathways involved in adult tissue homeostasis were identified, including regulation of proteostasis, cellular senescence and extracellular matrix remodelling. Putative effector genes of HF and ni-HF were enriched for proteoglycan pathways, including *ERBB2* the target of cancer therapies that is associated with impaired cardiac function^33,34^. Genes relating to the formation of aggresomes, cellular organelles store misfolded proteins for subsequent disposal throughautophagy^35^, were enriched, including *HSBP7* and *BAG3*. BAG3-mediated sarcomeric protein turnover is implicated as a mechanism underlying heart failure^36^. Pathways regulating cell division and senescence were identified, implicating tissue homeostasis and renewal in predisposition to age-associated organ dysfunction, Notably, these included the senescence-associated secretory phenotype, a distinct phenotype with a range of cell-autonomous and non-cell-autonomous effects that reinforce senescence. *IGFBP7*, a circulating anti-angiogenic factor produced by the kidney and the vasculature, was prioritised within a ni-HFpEF locus. IGFBP7 was recently reported to stimulate cardiomyocyte senescence and cardiac remodelling^37^. Pathways related to cardiac arrhythmia, including regulation of cardiac muscle contraction by calcium ion sensing and cell communication by electrical coupling were specifically enriched for ni-HF. The glucocorticoid-related pathways were enriched for all HF subtypes; glucocorticoids are primary stress hormones that are implicated in heart failure through both glucocorticoid and mineralocorticoid receptor-mediated effects^38^. Genes that were identified in the HF but not the non-ischaemic subtypes were enriched in pathways related to dyslipidaemia.

**Figure 3.**
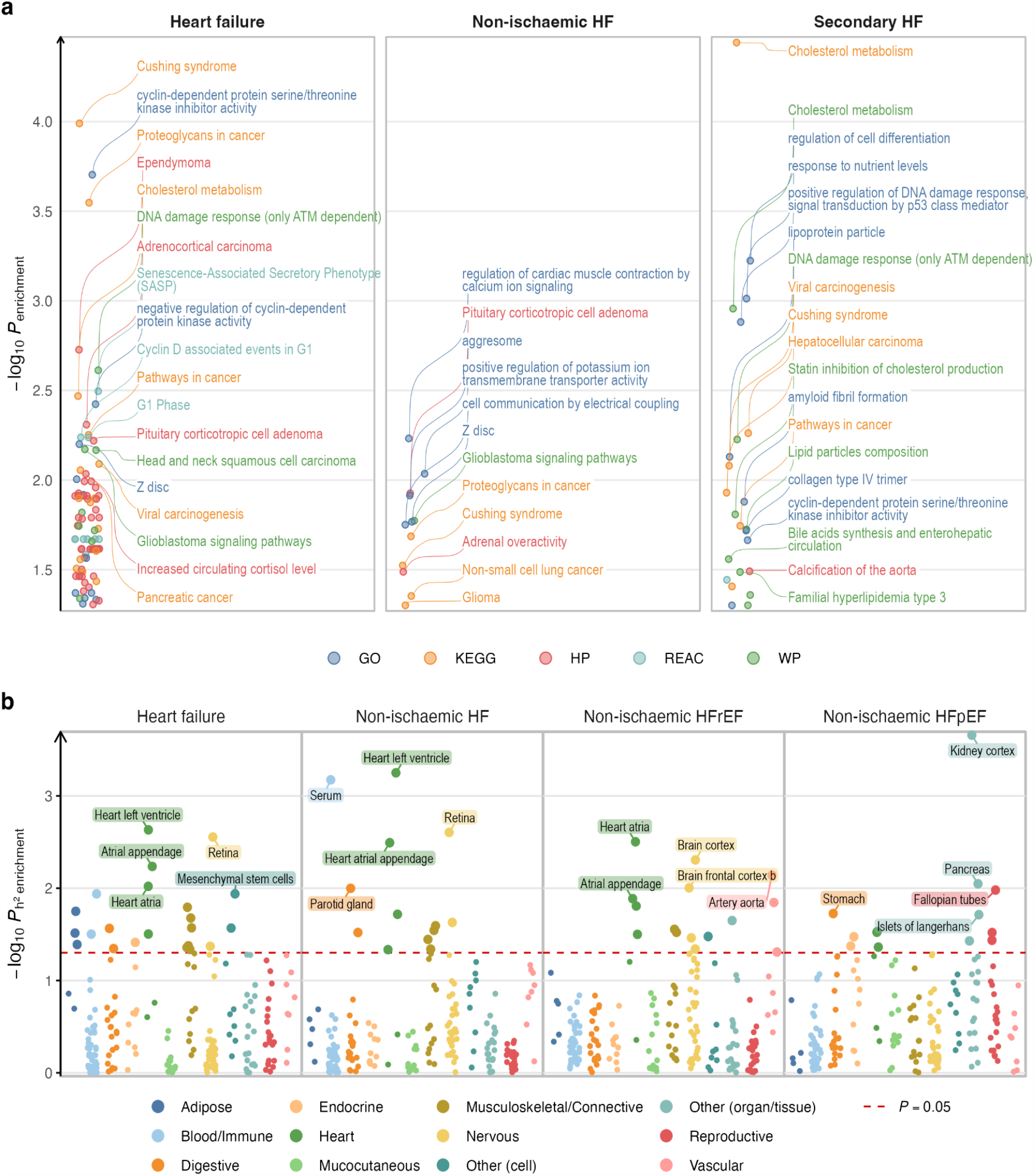
**a**. Enriched terms (Penrichment < 0.05) from Gene Ontology (GO), Kyoto Encyclopedia of Genes and Genomes (KEGG), Human Phenotype Ontology (HP), Reactome (REAC), and Wiki Pathways (WP). Up to top 20 enriched terms in each HF phenotype gene set are labelled. **b**. Heritability enrichment of 206 tissues and cell types, classified into 12 system / organ categories, across 4 HF phenotypes. Top 5 tissues / cell types per phenotype are labelled.

### A map of the organs, tissue and cells mediating heart failure risk

To identify tissues and cell types contributing to HF aetiology, we performed a heritability enrichment analysis across 206 tissues and cell types, representing 12 organs and systems, using sets of uniquely expressed genes (**Figure 3b, Supplementary Table 7**). 46 unique tissues and cell types were enriched (one-sided *P* < 0.05), highlighting the breadth of tissues and organ systems contributing to HF aetiology. Enrichment of cardiac tissues was the most frequent (15 enrichments), followed by musculoskeletal / connective tissues (12 enrichments), and nervous system tissues (8 enrichments). The most highly enriched tissues for HF, ni-HF, and ni-HFrEF, were cardiac whilst the kidney and pancreas were the most highly enriched for ni-HFpEF, highlighting the relative importance of these organs for specific HF phenotypes. Given the importance of cardiac tissues for ni-HFrEF, we extended the resolution of our enrichment analysis to individual cardiac cell populations, using single nucleus gene expression data from 16 non-failing human hearts^39^ (**Supplementary Figure 12a, Supplementary Table 8**). Cardiomyocytes were enriched for non-ischaemic HF and HFrEF subtypes, consistent with the pattern observed in the organ-level analysis.

To determine whether HF susceptibility genes were differentially regulated in failing versus non-failing hearts, we investigated cell-type specific expression profiles of the identified HF genes. 30/95 (30%) of ni-HF genes were differentially regulated in failing hearts, predominantly within fibroblasts and cardiomyocytes (**Supplementary Figure 12b-d, Supplementary Table 9**). Nuclear Receptor Subfamily 3 Group C Member 1 (*NR3C1*) which encodes glucocorticoid receptors was upregulated in 12 out of 14 cardiac cell types from failing hearts, consistent with the glucocorticoid pathway enrichment among the identified HF genes. These findings support the reported cardioprotective effect of glucocorticoid signalling via inhibition of pathological mineralocorticoid receptor signalling^40^ across human cardiac cell types, which may partially corroborate the unexplained beneficial effect of mineralocorticoid receptor antagonist drugs in HF^41^. Another prosurvival HF gene, *KLF12*, was upregulated in cardiomyocytes and cardiac fibroblasts. KLF12 is a transcription factor that is upstream of p53 and p21 (*CDKN1A*); methylation of the KFL12 transcription factor binding motif has been identified as a mechanism for *LMNA* cardiomyopathy ^42^. In contrast to prior reports, we did not observe differential expression of *IGFBP7* in cardiomyocytes from failing hearts ^37^.

### Graph modelling of phenome-wide pleiotropy identifies aetiologic clusters

To identify aetiological clusters underlying HF based on shared genetic aetiology, we tested the associations between sentinel variants at the 66 HF susceptibility loci and 294 diseases in UK Biobank, and modelled the associations using a pleiotropy network graph^43^. We found 207 pleiotropic associations at FDR <1% (corresponding to *P* < 0.001) among 46 / 66 (70%) HF loci and 79 / 294 (27%) unique phenotypes (**Supplementary Figures 13-14, Supplementary Table 10**). We constructed a HF pleiotropy graph network by representing loci and diseases as nodes and locus-phenotype associations as edges (**Supplementary Figure 15a**). Community detection analysis on the network revealed distinct 18 locus-disease aetiological clusters (**Figure 4, Supplementary Figure 15b, Supplementary Table 11**). The largest cluster, cluster 1, contained traits and diseases related to atherosclerosis and genes that were predominantly related to secondary HF loci. Cluster 2 comprised cardiac arrhythmias and dilated cardiomyopathy, and genes mostly associated with non-ischaemic HF. Cluster 4 centred on hypertension but contained genes for infection and cancer, while Cluster 5 comprised genes associated with non-ischaemic HF and ni-HFpEF, and disease including adiposity, diabetes, and carpal tunnel syndrome.

**Figure 4.**
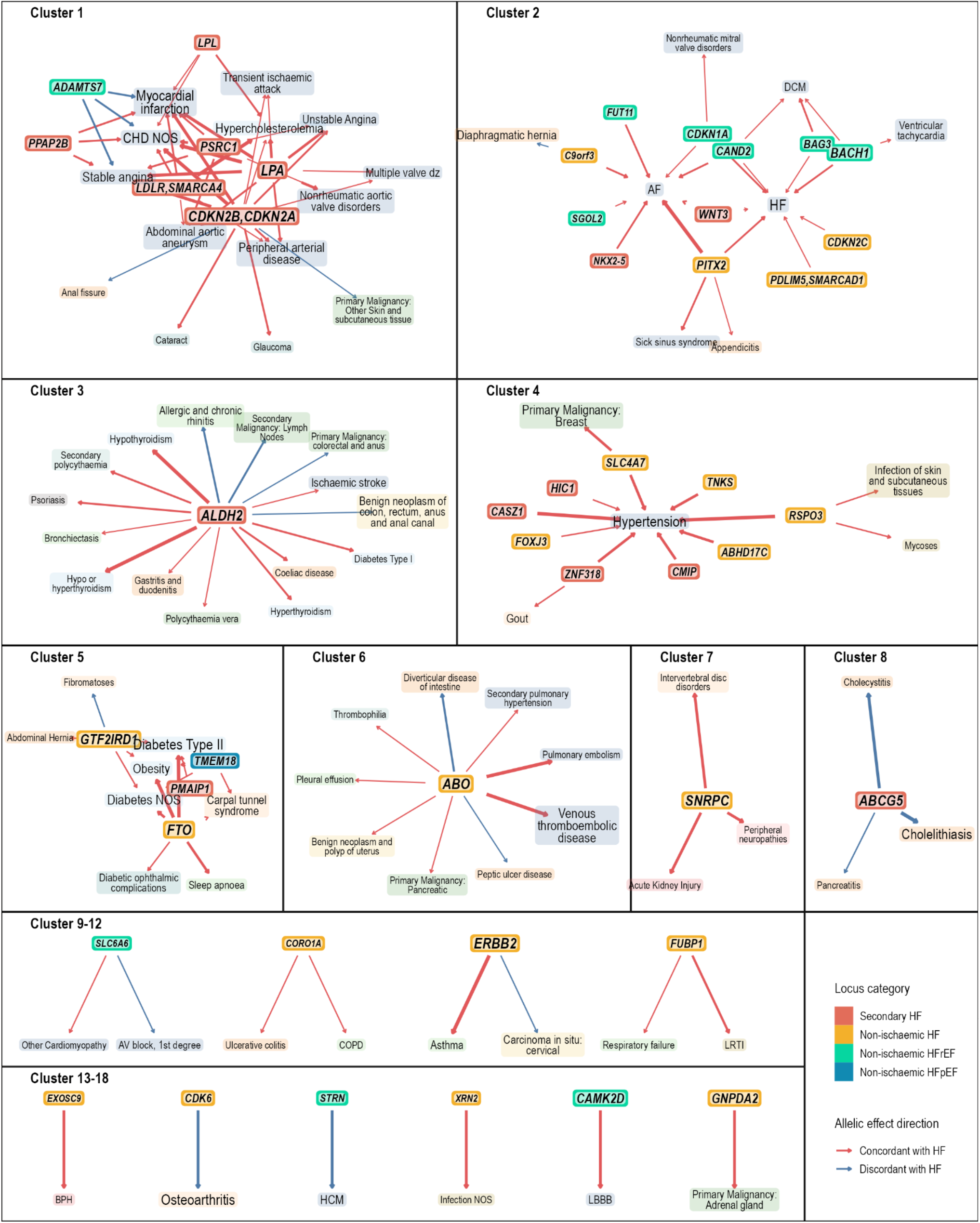
Aetiologic clusters identified through locus-phenotype pleiotropy network analysis of the phenome-wide association estimates. Nodes represent loci (solid background) and phenotype (translucent background), with size representing centrality measure. Edges represent locus-phenotype association, with thickness representing strength of association measured by absolute Z score.

Given the extensive pleiotropic effects of HF susceptibility loci, we further investigated the effects of sentinel variants in each locus on 24 GWAS traits representing major organ systems and image-based cardiac endophenotypes. We identified 270 genotype-phenotype associations at FDR <1%, of which 95 (35%) reached genome-wide significance in the original GWAS (**Supplementary Figure 16, Supplementary Table 12**). All tested traits were associated with at least one HF locus at FDR <1%, with coronary artery disease (CAD) and systolic blood pressure (SBP) showing the highest number (25 out of 66 loci / 38%). All lead variants in HF loci that were associated with DCM (6 loci) showed a concordant allelic effect (i.e. additional HF risk-increasing allele was associated with an increased disease risk), whereas all shared associations with HCM (14 loci) showed a discordant allelic effect, consistent with the opposite genetic effects on both traits ^44^. Colocalisation analysis revealed evidence of 105 common causal variants across 22 of the 24 traits tested (posterior probability > 0.8) at 42 out of 66 loci (**Figure 5a-c, Supplementary Table 14**), adding further evidence for pleiotropic associations and unveiling novel associations. For example, we found colocalisation of the *BACH1* ni-HFrEF locus with both coronary artery disease and dilated cardiomyopathy. *BACH1* is a transcription factor expressed in cardiac fibroblast and endothelial cells in response to mechanical stress and regulates YAP^45^. The BACH1-YAP transcriptional network exerts essential roles in both atherosclerosis and cardiac regeneration through the regulation of proliferative programs in cardiomyocytes^46^. Notably, six HF genes were found to colocalise with glomerular filtration rate (eGFR) or chronic kidney disease (CKD), including prefoldin subunit 1 (*PFDN1*) an important molecular chaperone for protein folding associated with mortality and cardiovascular phenotypes in mouse knockouts^47,48^.

**Figure 5.**
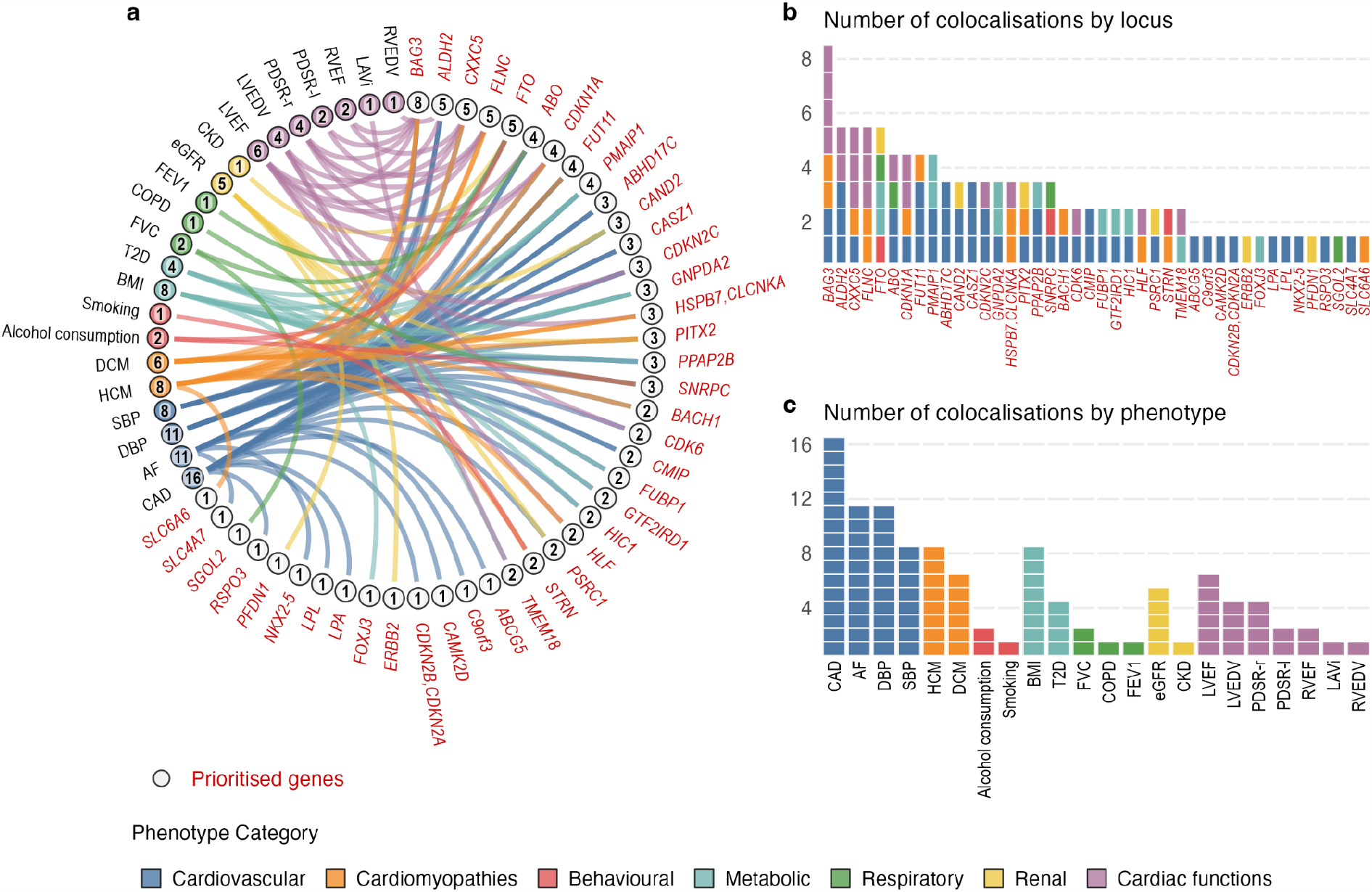
**a**. Colocalisations between HF and 22 GWAS phenotypes (out of 24 tested) across 42 (out of 66) loci with posterior probability of shared causal variants (*PP*_*coloc H4*_) > 0.8. Each band connects a pair of locus - phenotype nodes, and represents sharing of causal variants between the phenotype and HF at the locus. Annotated number represents total number of colocalisations by locus and phenotype, which is displayed in panel **b. & c**.

### Genetic appraisal of risk factors across the heart failure spectrum

Next, we sought to explore evidence for causal relationships between risk phenotypes associated with HF in the pleiotropy analysis, using genetic correlation and Mendelian randomisation (MR) analysis (**Figure 6, Supplementary Figure 17-19, Supplementary Tables 14-15**). First, we compared the estimated effects of upstream risk factors on HF and non-ischaemic HF. Trait associations were broadly similar between HF and ni-HF, except for coronary artery disease (CAD) for which there was evidence of causal effects on HF [MR odds ratio per doubling prevalence with inverse variance weighted estimator / OR_MR-IVW_ = 1.20, 95% confidence intervals = 1.18-1.22] but not on non-ischaemic HF [OR_MR-IVW_ = 1.02, 1.00-1.04]. We found evidence for the causal effects of systolic blood pressure (SBP), body mass index (BMI), and liability to atrial fibrillation (AF) across the HF phenotypic spectrum. SBP had the largest effect on ni-HFrEF [OR_MR-IVW_ = 1.92, 1.70-2.17 per standard deviation / SD] while BMI had the largest effects on ni-HFpEF [OR_MR-IVW_ = 1.87, 1.70-2.06 per SD]. Although type 2 diabetes and HF shared ∼23% genetic risk [squared genetic correlation / *r*_g_^2^ = 0.23], there was little evidence of a causal association between the two disorders [OR_MR-IVW_ = 1.04, 1.03-1.05; OR_MR-Egger_ = 0.98, 0.96-1.00 per doubling prevalence], suggesting that observational associations between T2D and HF largely reflect common upstream disease mechanisms, rather than direct causal effects of T2D on HF. Higher functional lung capacity (forced vital capacity, FVC; forced expiratory volume in 1 second, FEV1) was protective for HF [OR_MR-IVW_ = 0.93, 0.87-0.94 per SD FVC; OR_MR-IVW_ = 0.87, 0.83-0.92 per SD FEV1] and ni-HFpEF [OR_MR-IVW_ = 0.70, 95% CI = 0.61-0.80 per SD FVC; OR_MR-IVW_ = 0.86, 0.76-0.97 per SD FEV1]. We found evidence for effects of smoking behaviour on HF [OR_MR-IVW_ = 1.26, 95% CI = 1.19-1.33 per SD pack-years of smoking]. Positive associations of higher alcohol consumption with HF [OR_MR-IVW_ = 1.17, 95% CI = 1.06-1.29 per SD drinks / week] and ni-HFrEF [OR_MR-IVW_ = 1.87, 95% CI = 1.37-2.55 per SD drinks / week] were observed, consistent with clinical studies relating alcohol consumption to DCM^49^.

**Figure 6.**
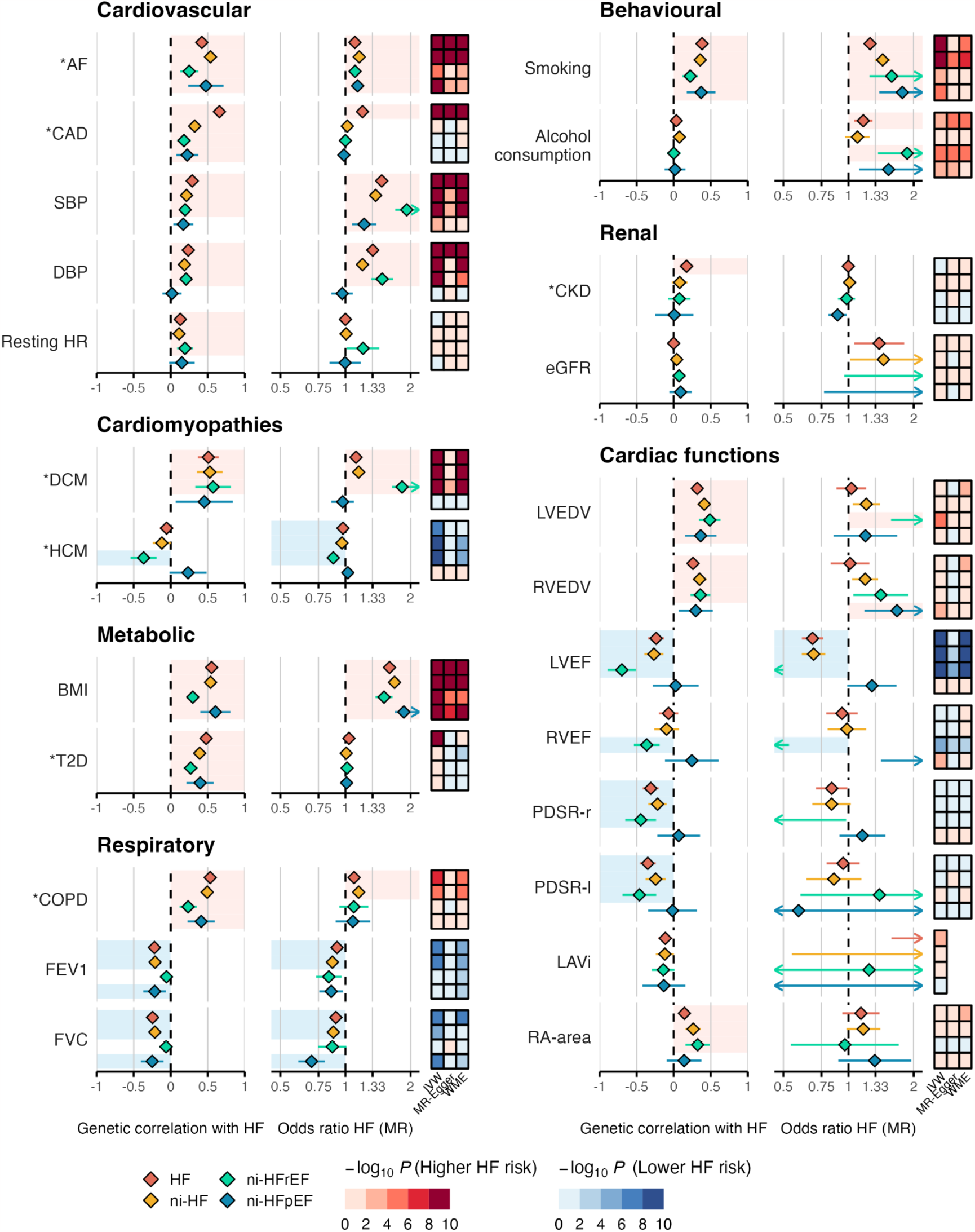
Bivariate genetic correlation (*r*_*g*_) and Mendelian randomisation (MR) estimates across 24 traits and 4 HF phenotypes. Asterisks (*) indicate binary traits. MR effects estimates are reported as odds ratio (OR_MR_) per doubling prevalence for binary traits; or per standard deviation increase for quantitative traits. Estimates which were robust to multiple testing adjustment and sensitivity analyses were indicated by light blue shade (for *r*_*g*_ < 0 and OR_MR_ < 1) or light red shade (for *r*_*g*_ > 0 and OR_MR_ > 1). The heat maps represent *P-values* for different MR models, colour-coded with direction of MR estimates and strength of associations.

Although renal tissues and traits (glomerular filtration rate, eGFR; chronic kidney disease, CKD) were associated with ni-HFpEF in the heritability enrichment (**Figure 4**) and colocalisation analyses (**Figure 6**), we found little or no evidence of genetic correlation [*r*_*g*_ = 0.17, SE = 0.05 between HF and CKD; and *r*_*g*_ = -0.001, SE = 0.052 between HF and eGFR]. These findings may reflect directional heterogeneity of locus-specific effects which results in a global genetic correlation. For example, there were 13 lead variants of HF associated with eGFR at FDR 1%, of which 8 had a concordant allelic effect and 5 had a discordant allelic effect on HF risk (**Supplementary Figure 16**). Paradoxically, we found suggestive evidence of a protective effect of CKD on ni-HFpEF [OR_MR-IVW_ = 0.89, 0.81-0.98] which may indicate confounding by competing diagnosis, whereby a CKD diagnosis reduces the likelihood of diagnosis with HF in patients presenting with fluid congestion^50^.

### The relationship between cardiac structure and function and heart failure

Traits and diseases associated with reduced contractile function were found to be associated with risk of all HF phenotypes, except ni-HFpEF (**Figure 6, Supplementary Figure 19**). As expected, the strongest effect was observed in ni-HFrEF [OR_MR-IVW_ = 0.18, 0.12-0.27 per SD LVEF; OR_MR-IVW_ = 1.83, 1.64-2.03 per doubling prevalence of DCM]. Conversely, traits associated with increased contractility were protective of ni-HFrEF [OR_MR-IVW_ = 0.88, 0.85-0.91 per doubling prevalence of HCM] and suggestive evidence that higher left ventricular ejection fraction is a risk factor for ni-HFpEF was observed [OR_MR-IVW_ = 1.28, 0.99-1.67 per SD LVEF]. These findings are consistent with divergent risk effects of subnormal and supranormal contractility DCM / HFrEF and HCM / HFpEF^44,51^. Despite the widely held hypothesis that diastolic dysfunction is the key driver for HFpEF, we found inconsistent evidence to support a causal association [OR_MR-IVW_ for ni-HFpEF = 1.16, 0.91-1.48 per SD PDSR-r].

## Discussion

The processes of evolution to animal life on land led to exquisite systems of body water regulation but an inherent susceptibility to systemic and pulmonary oedema^52^. Syndromes of fluid overload are a common clinical problem described since antiquity and are associated with elevated morbidity and mortality^53^. In the 20th century, cardiac dysfunction was identified as an important cause of fluid overload and formed the basis for the definition of heart failure. Increasingly, however, the importance of non-cardiac tissue in HF aetiology is emerging. By leveraging the de-confounding properties of germline genetic variation, we systematically appraise the aetiology of HF, highlighting key phenotypic traits, tissue, pathways and genes. Our findings generate new insights into the drivers of risk in particular HF subtypes, including for patients with preserved ejection fraction where therapeutic options are limited.

The insights from our study have several important translational implications. First, our appraisal of cardiovascular risk factor effects on HF subtypes yields several notable findings. Whilst clear effects of epicardial CAD and T2D on HF were observed, no effect on HF patients who had not undergone revascularisation or suffered from myocardial infarction was identified. Similarly, while T2D and HF share common genetic and environmental risk factors, we did not find strong evidence supporting a causal association between T2D and HF. These findings highlight the importance of managing upstream risk factors for diabetes and CAD, for the prevention of HF. SGLT2 inhibitors may exert their beneficial effects on HF risk and progression through modification of upstream physiology for both T2D and HF ^54^, such as higher body mass index; the effects of which are consistent across subtypes and may be independent of established risk factors^55^.

Second, our findings highlight the importance of cardiac contractility traits in HF. Non-ischaemic HFrEF was the most heritable subtype (h^2^_SNP_ = 11.8% ± 2.6%) and was highly correlated with DCM (*r*_g_ = 0.57 ± 0.12), as expected. We demonstrate opposing associations between contractility-related traits and ni-HFrEF and ni-HFpEF risk: higher baseline contractility decreased the risk of HFrEF, but likely increased the risk of HFpEF. Our findings do not provide positive evidence to support the widely held hypothesis that diastolic function is the major cause of HFpEF.

Third, our heritability enrichment analysis suggests that kidney, vascular, and metabolic tissues play a central role in the aetiology of HFpEF. The kidney is the primary organ for managing body fluid, and renal impairment is common in heart failure patients across the phenotypic spectrum. CKD, however, co-occurs more frequently with HFpEF than HF with mid-range or reduced ejection fraction^56^. It is possible that suggestive evidence of a paradoxical protective effect of CKD on ni-HFpEF reflects these diagnoses being alternative diagnoses for overlapping or indistinguishable clinical presentations. These findings motivate future studies to further investigate extra-cardiac drivers of fluid congestion in this patient population.

Fourth, our results reveal novel molecular mechanisms, both cardiac and extracardiac, across the heart failure spectrum. Tissue homeostasis emerges as a major mechanism underlying risk across the phenotypic spectrum of heart failure. Cell intrinsic mechanisms, including through modulation of *BACH1-YAP, KLF12-CDKN1A* activity, may influence myocardial tissue homeostasis through effects on cardiac growth. We found enrichment of the senescence-associated secretory phenotype components in pathway analysis and identify *IGFBP7* as a novel prioritised HFpEF gene, reflecting cell-autonomous effects of senescent cells on organismal aging^57^. *IGFBP7* is synthesised by the vasculature in response to tissue damage and may be an upstream driver of cell cycle arrest, fibrosis, and vascular rarefaction that are observed in HFpEF. *IGFBP7* is an established clinical biomarker of acute renal failure and may represent a target for novel therapeutics^37,58^.

In summary, by harnessing the de-confounding qualities of human germline variation, we map the aetiology of the heart failure spectrum, from organs to molecules. Our findings provide genetic evidence to bolster the concept of HFpEF as a multi-system disorder and yield insights into the biological mechanisms underlying subtypes of HF that may inform the development of more effective therapeutics to improve outcomes in those affected.

## Online Methods

### Study design

The present meta-analysis included 1,946,349 individuals from 42 studies participating in the HERMES Consortium (40 studies) or publicly released GWAS data of heart failure (Biobank Japan and FINNGEN r3). Sample genotyping was performed locally per study per phenotype per ancestry (one dataset) using high-density genotyping arrays and imputed against Haplotype Reference Consortium^59^ (75 datasets), 1000 Genomes Project^60^ (6 datasets), TOPMED^61^ (15 datasets), or population-specific reference panels (10 datasets).

Study-level GWAS was performed locally per dataset using logistic regression (for prevalent cases) or cox proportional hazard (for incident cases) assuming additive genetic effect with adjustment for sex, age at DNA draw, genetic principal components, and study-specific covariates. All participating studies were ethically approved by local institutional review boards and all study participants provided written informed consent. The meta-analysis was performed centrally at the coordinating centre in accordance with guidelines for study procedures provided by the University College London Research Ethics Committee. Details of study-level participant characteristics, genetic analysis, and cohort description are provided in **Supplementary Tables 16-18**.

### Phenotype definition

The present analysis investigated 4 phenotypes: 1) Heart failure (HF), 2) Non-ischaemic HF (ni-HF), 3) Non-ischaemic HFrEF (ni-HFrEF), and 4) Non-ischaemic HFpEF (ni-HFpEF). The HF phenotype includes any diagnosis of HF based on physician’s adjudication, hospital record review, or diagnosis codes. Non-ischaemic HF was defined by excluding antecedent ischaemic, valvular, and congenital heart diseases. Non-ischaemic HFrEF was defined as non-ischaemic HF with LVEF <50% based on cardiac imaging or diagnosis of left ventricular systolic dysfunction (LVSD) at any point. Non-ischaemic HFpEF was defined as non-ischaemic HF with LVEF ≥50% based on cardiac imaging without record of LVSD at any point. Phenotyping was performed separately in each participating study, using a harmonised multi-modal phenotyping algorithm (**Supplementary Note**) as a guide. Phenotype definition for FINGENN (release 3)^62^ and Biobank Japan^63^ follows the original study case definition. Details of study-level phenotype definition are provided in **Supplementary Table 19**.

### Study-level summary statistics quality control

GWAS summary statistics from each participating study were processed centrally with a quality control (QC) workflow implemented in *Snakemake*^*64*^ following the procedure described by Winkler T, *et al*^*65*^. Variants with more than two alleles, regression coefficients (log odds ratio or log hazard ratios) >10, standard error of the coefficients >10, minor allele frequency (MAF) <1%, imputation (INFO) score <0.6, or effective allele count (, calculated as 2 × *MAF* × (1 − *MAF*) × *N*_*sample*_ × *INFO*, of <50 were excluded from the analysis. Remaining variants with allele frequency difference >0.2 as compared against ancestry-specific reference panels were further excluded. The reference panels were derived from whole-genome sequence data from HRC and 1000 genomes project phase 3 version 5 (1000Gp3v5) for European ancestry, or 1000Gp3v5 for East Asian, South Asian, African, and Admixed American (Hispanic) ancestries. Genomic inflation adjustment was applied for summary statistics with genomic control coefficient (*λ*_*GC*_) > 1.1. In addition, the reported *P* values and quantile-quantile (QQ) plots from all studies were inspected to check for consistency and to identify spurious associations. Full details of the quality control procedure and results are provided in **Supplementary Materials**.

### Genome-wide association meta-analysis

Study-level GWAS results were meta-analysed using a fixed-effect inverse variance weighted model implemented in METAL^66^. For each phenotype, we calculated variant-level and summary-level effective sample size as 4 / (1/*N*_*case*_ + 1/*N*_*control*_) where *N*_*case*_ and *N*_*control*_ represent the number of cases and controls included in the meta-analysis. Variants with an estimated overall MAF <0.01, variant-level effective sample size < 10% of the meta-analysis effective sample size, or reported in only one study were further removed. The final results consist of genetic association estimates of 10,199,961 common genetic variants (MAF >1%) with HF; 9,414,975 variants with non-ischaemic HF; 9,198,919 variants with non-ischaemic HFrEF; and 8,277,415 variants with non-ischaemic HFpEF.

### Cross-ancestry allelic effect heterogeneity assessment

To account for a possible bias due to heterogeneity of allelic effect across ancestries, we performed a sensitivity analysis using a meta-regression technique to model ancestry-specific allelic effect by incorporating axes of genetic variation derived from genome-wide metrics of diversity between populations, as implemented in MR-MEGA^67^. Using this technique, we also estimated overall *P* value for associations accounting for allelic effect heterogeneity across ancestries, *P* value for heterogeneity correlated with ancestry, and *P* value for residual heterogeneity. Concordance between *P* values from the fixed-effect meta-analysis with METAL and those from MR-MEGA were checked by calculating Pearson’s correlation coefficient and plotting the two vectors on a two-dimensional plane across conditionally independent variants. Given the limited non-European ancestry sample for non-ischaemic HF subtypes, this comparison was only performed using the overall HF phenotype.

### Identification of genomic susceptibility loci across HF subtypes

Genomic susceptibility loci for HF were identified using sets of conditionally independent variants identified through a chromosome-wide stepwise conditional-joint analysis using Genome-wide Complex Trait Analysis (GCTA) software^68^. Conditionally independent variants across 4 HF phenotypes with joint *P* value < 5 x 10^-8^ that are physically located within 500 kilobases of each other were aggregated into one set. A genomic locus was then defined as the genomic region within 500 kilobases upstream and downstream of the farthest variants of each aggregated set of conditionally independent variants.

The identified genomic loci were labelled with an incremental one-based integer sequence based on phenotype order (HF, ni-HF, ni-HFrEF, and ni-HFpEF), chromosome, and base pair positions. A genomic locus was declared as novel if all conditionally independent variants within the locus and any of the sentinel variants reported at *P* < 5 x 10^8^ in previous GWAS of HF^1–5^ were physically located more than 250 kb away and not in LD (*R*^2^ < 0.2). To model LD between genetic variants, we used a reference panel derived from individual-level imputed genotype data of a randomly sampled 10,000 UK Biobank participants with an ancestry composition proportionally matched to the present meta-analysis (Hispanic ancestry was not included due to unavailability in UK Biobank and small proportion in the overall sample).

Further, association between a locus and a HF subtype was categorised based on *P* values at cut-off values of *P* < 5 x 10^-8^ (genome-wide significant), *P* < 0.05 / number of identified loci (replicated at Bonferroni-adjusted threshold), *P* < 0.05 (nominally significant), and *P* ≥ 0.05 (no evidence of association). For presentation, loci associated with any non-ischaemic HF subtype at Bonferroni-adjusted or genome-wide significance thresholds were labelled as non-ischaemic HF loci, and the remainings were labelled as secondary HF loci. Non-ischaemic HF loci that are associated with ni-HFrEF at Bonferroni-adjusted or genome-wide significance thresholds were labelled as ni-HFrEF loci, and the remainings were labelled as ni-HFpEF loci.

### Genetic architecture assessment

To assess genetic architecture and polygenicity across HF phenotypes, we compared quantiles of the expected and observed genome-wide genetic association *P*-values using quantile-quantile (QQ plot) and calculated genomic control coefficient (*λ*_*GC*_). An early inflation on QQ plot with *λ*_*GC*_ > 1.1 suggests genome-wide genomic inflation, which may be due to polygenic genetic architecture or confounding by population stratification. To distinguish between the two, we calculated *λ*_*GC*_ assuming 1000 participants (*λ*_*GC*-1000_), and estimated LD score regression slope using LDSC software^69^, whereby values close to 1 indicate that any observed genomic inflation is likely due to polygenic genetic architecture. To minimise confounding by population structure, LDSC regression was performed using European ancestry meta-analysis subset and LD reference panel. We investigated the distribution of allelic effect on HF across the spectrum of allele frequency by plotting the risk ratio per additional minor allele as a function of minor allele frequency of conditionally independent variants. To assess the shape of the relationship, we fitted two separate local polynomial regression with locally estimated scatterplot smoothing (LOESS) for groups of variants associated with increased and decreased risk of HF. To increase precision of the fitted regression, a sub-genome-wide threshold association at a false discovery rate (FDR) <1%, estimated using the *qvalue* package in R^70^, was used to identify conditionally independent variants.

### SNP-based heritability estimation

The proportion of variance in HF risk explained by common SNPs, i.e. SNP-based heritability 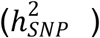, was estimated from GWAS meta-analysis summary statistics using Linkage-Disequilibrium Adjusted Kinships (LDAK) SumHer software^71^ with LDAK-Thin and BLD-LDAK heritability models^72^. The 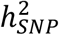 estimates were calculated on a liability scale, which assumes that a binary phenotype has an underlying continuous liability, and above a certain liability threshold an individual becomes affected^73^. To obtain a more accurate estimate, we used the European ancestry meta-analysis summary statistics with pre-computed tagging files derived from 2,000 individuals White British individuals, and used a correction for sample prevalence by calculating the effective sample size assuming an equal number of cases and controls^74^. The conversion to liability scale was calculated using population prevalence derived from the total number of cases divided by the total number of cases and controls included in the meta-analysis.

### Polygenic risk score analysis

To explore the potential utility of SNP-based heritability for prediction, we explored the association between polygenic score of HF (PGS_HF_) and HF risk in 346,667 UKB European participants, including 13,824 prevalent and incident HF cases. The PGS_HF_ was constructed as a weighted sum of allelic count of 1,012,059 genetic variants selected using the LDpred2-auto model^75^, with weights derived from the present GWAS meta-analysis of HF excluding UK Biobank. Odds ratio of HF per standard deviation of PGS_HF_ was estimated using logistic regression with binary HF status as response variable, standardised PGS_HF_ as predictor, and sex, age and first 10 ten genetic principal components (PC)s as covariates. To estimate risks of HF in individuals with high PGS_HF_, we grouped participants into deciles of PGS_HF_, and calculated odds ratios of the top decile as compared against the fifth and the first (bottom) decile using logistic regression.

### Fine mapping of causal variants and functional assessment

We performed functionally-informed fine-mapping using PolyFun^7^ and SuSiE^76^ to prioritise likely causal variants in each identified genomic locus for HF. We used precomputed functionally-informed prior causal probabilities of 19 million imputed UK Biobank SNPs with MAF>0.1%, based on a meta-analysis of 15 UK Biobank traits^7^ and genome-wide association estimates from the current analysis to calculate prior causal-probability proportional to per-SNP heritability of phenotype under analysis. The resulting estimates were passed on to the sum-of-single-effects fine-mapping model implemented in SuSiE, assuming at most 5 causal variants per locus, to calculate per-SNP posterior inclusion probability (PIP) and to construct 95% credible sets of likely causal variants. To minimise bias due to differential ancestry composition, we used effect estimates from meta-analysis of European ancestry GWAS and LD reference panel from 10,000 randomly selected UK Biobank participants of European ancestry. To assess functions and consequences of fine-mapped variants, we extracted variant-level information on nearest gene(s), genic functions, and Combined Annotation-Dependent Depletion (CADD)^8^ Phred score from ANNOVAR^77^ and OpenTargets Genetics^12^.

### Prioritisation of effector genes for HF

To identify effector genes for HF, we implemented a two-step in silico gene prioritisation approach. In step 1, we identified *candidate* gene set using a combination of three *predictors*:

1. Polygenic priority score (PoPS) Polygenic enrichment of gene features derived from cell-type specific gene expression, biological pathways, and protein-protein interactions^13^
2. Variant to gene score (V2G) Highest V2G score across variants in the 95% credible set and variants in high LD (r^2^ >0.8) with the conditionally independent variants within the locus. The V2G score was extracted from the OpenTargets Genetics^12,78^, and was derived from in silico functional prediction, expression and protein quantitative trait loci, chromatin interactions, and distance to gene’s canonical transcription start site portal
3. Transcriptome-wide association study (TWAS) *P* value for association of overall predicted gene expression across tissues with HF calculated using S-MulTiXcan^14^ with GTEx^79^ v8 MASHR models.

Genes with highest PoPS, highest V2G score, or lowest TWAS *P* value within a locus were considered as *candidate* effector genes for HF. In step 2, we prioritised these genes using 3 boolean (True / False) *classifiers*:

1. Top ≥2 Whether a gene ranked top by 2 of the 3 *predictors* (PoPS, V2G, TWAS)
2. Mendel Whether a gene is associated with at least one Mendelian disease term that is enriched at FDR <1% as estimated from Mendelian gene set enrichment analysis using MendelVar^16^.
3. Coloc Whether gene expression level in tissue with lowest *P*-value from the multi-tissue TWAS analysis colocalised with at least 1 HF phenotype under study (posterior probability of shared causal variants >0.8). The colocalization analysis was performed with the R *coloc* package allowing for multiple causal variants^15^ using gene expression data from GTEx v8^79^.

In addition, we derived an *Overall* predictor score based on weighted average of PoPS, highest V2G, and -log_10_ *P* MulTiXcan values with 2:2:1 weight ratio, scaled to 0-100 value using a quantile transformation with uniform output distribution as implemented in python *scikit-learn*^*80*^ library. Finally, for each locus, we ranked genes based on total classifier score (sum of *True* values) and the overall predictor score. Genes that are top-ranked or have a total classifier score ≥2 were *prioritised* as most likely effector genes for HF.

Analyses which require GWAS summary statistics (PoPS, TWAS, and Coloc) were performed separately for each of the 4 HF phenotypes using meta-analysis results from the European subset. Locus-specific results for gene prioritisation were extracted from the overall HF phenotype for loci with *P*_HF-GWAS_ < 5 x 10^-8^ (locus 1-56), or from HF phenotype with the lowest *P*_*GWAS*_ < 5 x 10^-8^ for subtype-specific loci (ni-HF for locus 57-61, ni-HFrEF for locus 62-64, ni-HFpEF lor locus 65-66).

### Heritability enrichment analysis

To identify relevant tissues and cell types involved in HF pathology, we estimated heritability enrichment of specifically expressed genes using LDSC-SEG^81^. We used specifically expressed gene sets for 206 tissues and cell types from GTEx and Franke Lab^82,83^, classified into 12 organ / system categories (**Supplementary Table 20**). For each set, LDSC-SEG tested the enrichment for per-SNP heritability attributed to the given set, conditional on the set that includes all genes and a baseline model consisting of 52 genomic annotations (including genic regions, enhancer regions and conserved regions) using stratified LDSC regression framework^84^. A positive regression coefficient, statistically tested using a one-sided *P*-value test, represents a positive contribution of a given tissue / cell type to trait heritability. For this analysis, we used the European ancestry meta-analysis subset with pre-computed LD score weights derived from the 1000 Genomes European reference panel^84^.

Further, based on the observation that heart tissues are the major contributors to SNP heritability in almost all HF phenotypes, we extended the heritability enrichment analysis to 15 cell types derived from single-nucleus RNA sequencing (snRNA-seq) experiment on 185,185 cell nuclei from 16 non-failing human heart donors^39^ (**Supplementary Table 21**). To identify specifically expressed genes for each cell type, we followed the approach described in Finucane et al. (2018)^81^ by taking top 10% genes with the highest *t*-statistic for specific expression in a given cell type (compared to other cell types). The sets of specifically expressed genes for each cardiac cell type were used to perform the heritability enrichment analysis with LDSC-SEG as described above.

### Single-nucleus differential gene expression in failing vs. non-failing heart

To assess the extent to which transcriptional pattern of risk genes for HF changes in failing heart, we performed single-nucleus differential gene expression analysis of candidate effector genes within HF loci by comparing transcription level in cell nuclei from non-failing heart as described above with cell nuclei from 28 failing heart donors diagnosed with end-stage dilated cardiomyopathy (DCM) or hypertrophic cardiomyopathy (HCM)^39^.

Samples were collected from Myocardial Applied Genetics Network (MAGNet; www.med.upenn.edu/magnet), with snRNA-seq processed using CellBender^85^ and Cell Ranger^86^ as previously described^39,87,88^. Where available, for each gene in a given cell type, we computed statistics for differential gene expression between failing and non-failing samples using limma–voom^89,90^ model adjusting for age and sex. To account for the correlation in expression among nuclei from a given individual, we summed counts for genes across nuclei for each patient within each cell type, requiring a minimum of 25 nuclei. We excluded mitochondrial genes and ribosomal genes, removed genes in less than 1% of nuclei in the given cell type of both groups being compared, and applied an additional filter for lowly expressed genes using the filterByExpr(group=group) function. Two-sided P-values were calculated and A Benjamini–Hochberg correction was applied for multiple testing correction.

A gene was declared to be differentially expressed if it survived multiple-testing correction at FDR-adjusted P-value <0.01 and showed a concordant differential expression sign in both CellBender and Cell Ranger quantifications and had no background contamination as estimated in CellBender. To test whether differentially expressed genes in a given cell type are overrepresented amongst prioritised HF genes, we performed a Fisher exact test implemented in *R*, using all genes tested in the given cell type as the background set.

### Pathway enrichment analysis

To identify biological pathways and processes that are relevant to HF pathology, we tested for overrepresentation of biological terms in prioritised genes for HF using g:Profiler^91^. To account for aetiological differences underlying HF subtypes and uncertainty in gene prioritisation, we tested 6 gene sets constructed from combinations of 3 phenotypic classifications of loci (all loci, secondary HF loci, non-ischaemic HF loci); and 2 gene prioritisation categories (*Candidate* gene set: genes that are ranked top by PoPS, TWAS, or V2G predictor and *Prioritised* gene set: candidate genes with classifier score ≥2 or highest overall predictor score). For each gene set, we performed an unordered enrichment analysis of biological terms in Kyoto Encyclopedia of Genes and Genomes (KEGG)^92^, Reactome (REAC)^93^; Wiki Pathways (WP)^94^; and Gene Ontology (GO)^95,96^. For presentation (**Figure 3a**), we excluded terms with more than 2,000 genes (representing ∼10% of protein coding genes in the human genome) and included terms with adjusted *P* < 0.05 following g:Profiler correction for multiple testing. GO terms were summarised by highlighting only driver terms as identified by the g:Profiler clustering algorithm. Terms that are enriched in both *candidate* and *prioritised* gene sets were collapsed by presenting the median enrichment *P* value on -log_10_ scale.

### Phenome-wide association studies in UK Biobank

To characterise pleiotropic effects of HF genetic loci across human diseases, we performed a phenome-wide association study (PheWAS) by testing the associations of the lead variants at 66 HF loci with 294 disease phenotypes in 408,480 UK Biobank participants of European ancestries. Phenotypes were derived from clinical events recorded in data from linked hospital admission, death certificate, primary care visit, and self-reported cancer diagnosis, non-cancer diagnosis, and procedure history; using expert-curated phenotype definition and category described elsewhere^43^. For a given phenotype, participants were classified as cases if they ever had a record of at least one matching code in any data source; otherwise as controls. For each phenotype - lead variant pair, we ran a case-control genetic association analysis implemented in PLINK2, requiring at least 100 cases and 100 controls per phenotype. The association analysis was performed using a logistic regression model assuming an additive allelic effect and adjusted for sex, genotyping array, and first ten genetic principal components. For presentation, allelic effect estimates were aligned to represent the effect of one additional HF risk-increasing allele.

### Pleiotropy network analysis

Using results from the identified genotype-phenotype associations from the locus PheWAS, we performed a network analysis to evaluate the connections between HF genetic loci based on their pleiotropic effects across human diseases. First, we constructed a network dataset with nodes (vertices) representing loci or phenotypes; and edges representing unidirectional associations of the locus lead variant with a given phenotype. The resulting network was then visualised using the Davidson-Harel layout algorithm^97^. To help with interpretation, edges on the graph were coloured by the effect direction of HF risk-increasing allele, with thickness of the line representing absolute Z statistics for the association. Phenotype nodes were labelled by disease name and coloured by disease category; whereas locus nodes were labelled by most likely effector genes. Eigenvector centrality measure was calculated to estimate node influence within the network (**Supplementary Figure 20, Supplementary Table 22**), and then mapped to the node size so that a more influential node appears larger.

Further, we performed a data-driven community detection analysis on the graph using a walktrap algorithm^98^ weighted by absolute Z score of the edges to identify clusters of densely connected nodes. Overall modularity^99^ of the resulting clusters was calculated to quantify the strength of the division, where a higher value reflects a connection between nodes that is denser for nodes within the same cluster but sparser for nodes belonging to different clusters. The network analysis and visualisation was performed using the *igraph, tidygraph* and *ggraph* package in R.

### Locus-specific pleiotropy assessment

We investigated the local pleiotropic effects of sentinel variants in 66 genetic loci on 24 HF-relevant GWAS traits (**Supplementary Table 23**). Sentinel variants were defined as conditionally independent variants showing lowest *P*-value for association with HF phenotypes within a locus. For loci associated with both all-comer HF and at least one non-ischaemic HF phenotype, sentinel variants were taken from all-comer HF. Estimates for sentinel variant that were not reported in a target GWAS were substituted with those of a proxy variant with highest LD *r*^2^ (requiring at least LD *r*^2^ >0.8), identified using *plink*^*100*^ with LD reference panel derived from 10,000 random samples of UK Biobank European participants. For presentation (**Supplementary Figure 15**), effect estimates were converted to Z-scores derived from the regression coefficients (betas) divided by their standard errors, with direction of effect aligned to reflect the additive effect of HF-risk-increasing allele. Loci and GWAS traits were ordered using hierarchical agglomerative clustering with average linkage method, implemented with the *hclust* function in R 4.2.0. Distances between loci / traits for clustering were calculated with the Euclidean method using vectors of absolute Z-scores (reflecting allele-agnostic effects across tested loci / traits).

### Cross-trait colocalisation

To test whether locus-specific genetic associations with HF and a given trait derived from common causal variants, we performed pairwise cross-trait colocalisation analysis implemented in R *coloc* package^15^, allowing multiple causal variants per locus by integrating Sum of Single Effects (SuSiE) regression framework^76^. This approach first runs SuSiE to detect credible set(s) *L*_*1*_ and *L*_*2*_ corresponding to trait 1 and trait 2 respectively; followed by colocalisation to estimate posterior probabilities of common causal variants (hypothesis 4, *H*_*4*_) for each pairwise combination of elements of *L*_*1*_ x *L*_*2*_^*15*^. To run *coloc*, we used the default marginal prior probabilities for association with trait 1 (*p*_*1*_) and trait 2 (*p*_*2*_) of 10^-4^, and a prior probability of joint causal association (*p*_*12*_) of 10^-5^. A posterior probability of *H*_*4*_ (*PP*_*coloc H4*_) >0.8 in at least one pair of credible sets was considered as evidence of shared causal variants. The colocalisation analysis was performed for each identified HF locus using an overlapping set of variants available in both GWAS of HF and trait under analysis situated within the locus. Genetic association estimates for HF were extracted from GWAS of the overall HF phenotype for loci with *P*_HF_ < 5 x 10^-8^ (locus 1-56); or otherwise from GWAS of non-ischaemic HF phenotypes for subtype-specific loci: ni-HF for locus 57-61, ni-HFrEF for locus 62-64, and ni-HFpEF for locus 65-66.

### Genetic correlation

To assess the extent to which genetic associations are shared amongst HF subtypes, we performed genetic correlation analysis across pairs of 4 HF subtypes under analysis, and genetic correlation between pairwise combinations of each HF subtype and 24 HF-related GWAS traits. The genetic correlation analysis was performed using bivariate linkage disequilibrium score (LDSC) regression^101^ with input GWAS summary statistics taken from the present study (for HF phenotypes) and publicly available data (for other traits). Genetic correlation of HF and other binary traits was estimated on a liability scale, with sample prevalence estimated as the number of cases divided by total number of samples in the GWAS and population prevalence taken from literatures if available, or assumed to equate the sample prevalence otherwise (**Supplementary Table 23**).

### Mendelian randomisation

We estimated the causal effects of each of the 24 HF-related traits (**Supplementary Table 23**) on each of the 4 HF subtypes analysed in the present study using two-sample Mendelian randomization (MR) as implemented in the *MendelianRandomization* package^102^. MR instruments for the exposure traits (phenotypes other than HF) were selected from genetic variants available in both GWAS of exposure and outcome (HF phenotypes) traits using LD-based clumping algorithm with *P* value threshold of 5 x 10^-8^ and LD *r* ^2^ threshold of 0.05 implemented in PLINK^100^. For each exposure trait, we estimated the causal association with each HF phenotype using inverse-variance weighted (IVW) MR estimator; and performed sensitivity analyses with MR-Egger and weighted median estimators (WME)^103,104^. Traits that survived the multiple testing adjustment at FDR <1% in IVW analysis and showed consistent direction of effect in sensitivity analyses were considered causal.

### Multiple testing adjustment

Where mentioned in the text, we performed multiple testing adjustments by controlling false discovery rate (FDR) using the Benjamini-Hochberg procedure^105^ as implemented in the *p*.*adjust(method = “BH”)* function in *R*. For each analysis, adjusted *P*-values less than a tolerable type-I error rate (alpha) of 0.01, corresponding to false discovery rate of 1%, were considered to survive the multiple testing correction.

## Data availability

GWAS summary statistics from the meta-analysis will be made available on the Cardiovascular Disease Knowledge Portal (https://cvd.hugeamp.org/) and GWAS Catalog (https://www.ebi.ac.uk/gwas/summary-statistics) upon publication.

## Code availability

A sample code to define heart failure phenotypes in UK Biobank is available on: https://github.com/ihi-comp-med/ukb-hf-phenotyping. Other codes to perform key analyses presented in this work will be made available on https://github.com/ihi-comp-med/hermes2-gwas

## Acknowledgements

A.Henry was supported by the BHF Cardiovascular Biomedicine PhD studentship (FS/18/65/34186). R.T.L. and A.Henry are partly supported by a Pfizer Innovative Targets Exploration Network Grant. The project was additionally supported by BigData@Heart Consortium funded by the Innovative Medicines Initiative-2 Joint Undertaking under grant agreement No. 116074, the UCL British Heart Foundation Accelerator (AA/18/6/34223), National Institute for Health Research University College London Hospitals Biomedical Research Centre (NIHR203328), and Health Data Research UK (MR/S003754/1). Analyses using the UK Biobank resource presented in this work were conducted under Application Numbers 9922, 15422, 12113, and 47602. The authors thank all research participants included in the presented work. The views expressed in this work are those of the authors and not necessarily those of the funders. Additional study-level acknowledgements are provided in **Supplementary Table 18**.

## Author contributions

*Conceptualisation:* R.S.V., J.G.S., H.Holm, Sonia Shah, P.T.E., A.D.H., Q.W., R.T.L.

*Methodology:* A.Henry, M.D.C., D.S., Sonia Shah

*Project administration:* A.Henry, H.I., D.M., R.T.L.

*Formal analysis:* A.Henry, X.M.

*Software:* A.Henry, C.F., D.S., S.C., A.F.S.

*Data Curation:* A.Henry, H.I.

*Writing - Original Draft:* A.Henry, R.T.L.

*Visualisation:* A.Henry

*Resources:* C.F., M.D.C., S.D., J.Gratton, S.C., J.G.S., H.Holm, P.T.E., A.D.H.

*Investigation:* M.D.C., I.B., C.R., D.M.

*Supervision:* F.W.A., T.P.C., M.P.D., M.E.D., C.C.L., N.J.S., R.S.V., J.G.S., H.Holm, Sonia Shah, P.T.E., A.D.H., Q.W., R.T.L.

*Funding acquisition: R*.*T*.*L*. F.W.A., T.P.C., M.P.D., M.E.D., C.C.L., N.J.S., Svati Shah, R.S.V., J.G.S., H.Holm, P.T.E.,

A.D.H., Q.W., R.T.L. are members of the HERMES Executive Committee who provided additional supervision of the work.

Other co-authors not listed above contributed to data generation, funding acquisition, formal analysis, and supervision at individual study level.

Lists of contributors from Genes & Health Research Team, DBDS Genomic Consortium, and HERMES Consortium are provided in **Supplementary Note**.

All authors have reviewed and approved the final version of the manuscript.

## Competing interests

A.Henry and R.T.L. received funding from Pfizer. J.S.W. have acted as a consultant for MyoKardia, Pfizer, Foresite Labs, and Health Lumen, and received institutional support from Bristol-Myers Squibb and Pfizer. S.d.D. was supported through grants from AstraZeneca, Roche Molecular Science/DalCor. J.R.K. declares stock ownership in AbbVie, Abbott, Bristol Myers Squibb, Johnson & Johnson, Medtronic, Merck, Pfizer. N.A.M. received speaking honoraria from Amgen and is involved in clinical trials with Ionis, Amgen, Pfizer, and Novartis. B.M.P. serves on the Steering Committee of the Yale Open Data Access Project funded by Johnson & Johnson. C.T.R. received honoraria for scientific advisory boards and consulting from Anthos, Bayer, Bristol Myers Squibb, Daiichi Sankyo, Janssen, Pfizer, and received institutional research grants from Anthos, AstraZeneca, Daiichi Sankyo, Janssen and Novartis. M.S.S. received significant research grant support from Abbott Laboratories, Amgen, AstraZeneca, Bayer, Critical Diagnostics, Daiichi-Sankyo, Eisai, Genzyme, Gilead, GlaxoSmithKline, Intarcia, Janssen Research and Development, The Medicines Company, MedImmune, Merck, Novartis, Poxel, Pfizer, Quark Pharmaceuticals, Roche Diagnostics, and Takeda; and has received consulting fees from Alnylam, AstraZeneca, Bristol-Myers Squibb, CVS, Amgen. A.A.V. received consultancy fees and/or research support from AnaCardia, AstraZeneca, Bayer, BMS, Boehringer Ingelheim, Corteria, Cytokinetics, EliLilly, Moderna, Novartis, NovoNordisk, Roche Diagnostics. M-P.D. declares holding equity in Dalcor Pharmaceuticals, unrelated to this work.

Members of the TIMI Study Group (ENGAGE, FOURIER, PEGASUS, SAVOR, SOLID) have received institutional research grant support through Brigham and Women’s Hospital from: Abbott, Amgen, Anthos Therapeutics, ARCA Biopharma, Inc., AstraZeneca, Bayer HealthCare Pharmaceuticals, Inc., Daiichi-Sankyo, Eisai, Intarcia, Ionis Pharmaceuticals, Inc., Janssen Research and Development, LLC, MedImmune, Merck, Novartis, Pfizer, Quark Pharmaceuticals, Regeneron Pharmaceuticals, Inc., Roche, Siemens Healthcare Diagnostics, Inc., Softcell Medical Limited, The Medicines Company, Zora Biosciences, Caremark, Dyrnamix, Esperon, IFM Pharmaceuticals, MyoKardia.

The authors who are affiliated with deCODE genetics/Amgen Inc. and the authors affiliated with Pfizer Inc. declare competing financial interests as employees.

The remaining authors declare no competing interests.

## Notes

### Author Declarations

Ethics committee of University College London gave ethical approval for this work

