## Supplementary Figures 1-20 for "Mapping the aetiological foundations of the heart failure spectrum using human genetics"

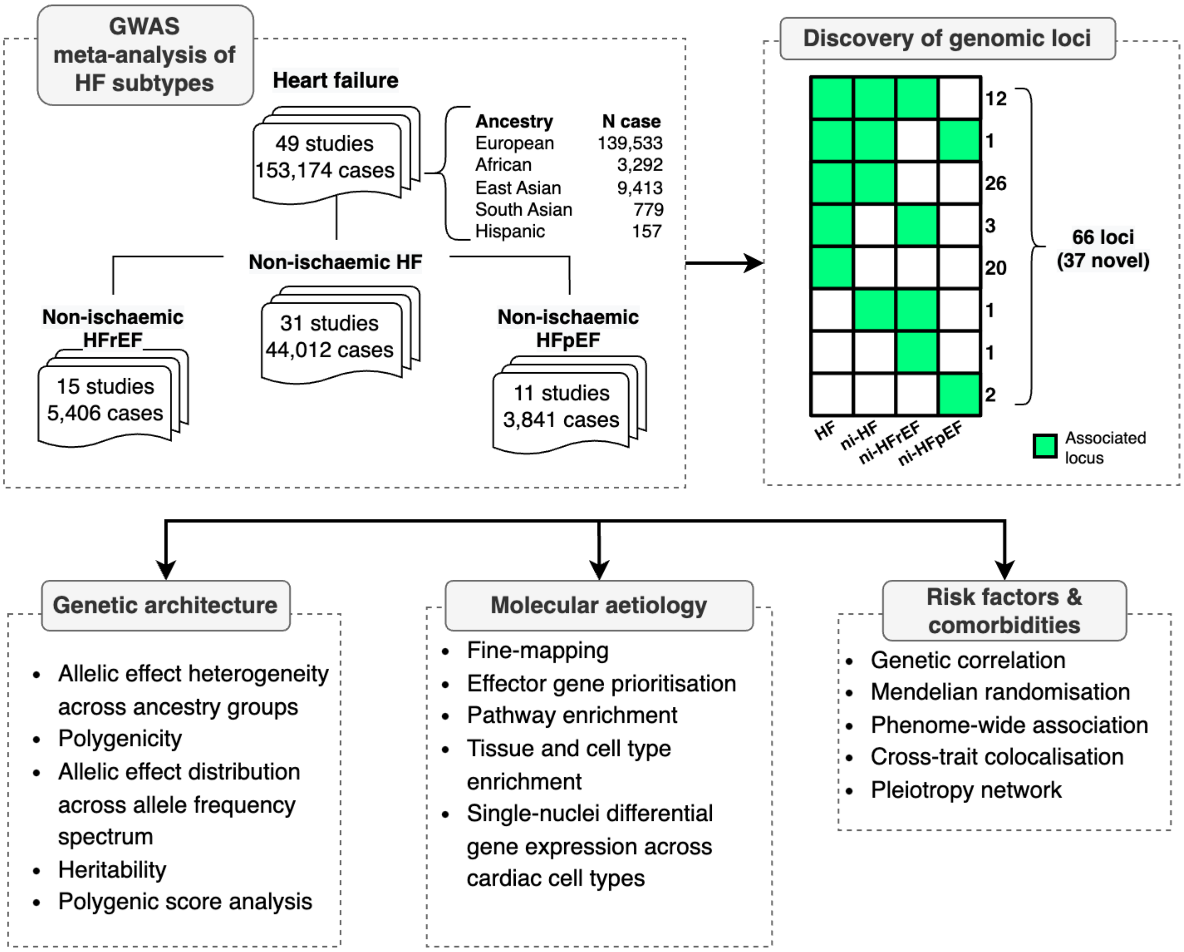


**Supplementary Figure 1.** Study design. GWAS: genome-wide association study, HF: heart failure, HFrEF: heart failure with reduced ejection fraction, HFpEF: heart failure with preserved ejection fraction, ni-HF: non-ischaemic heart failure, ni-HFrEF: non-ischaemic heart failure with reduced ejection fraction, ni-HFrEF: non-ischaemic heart failure with preserved ejection fraction.

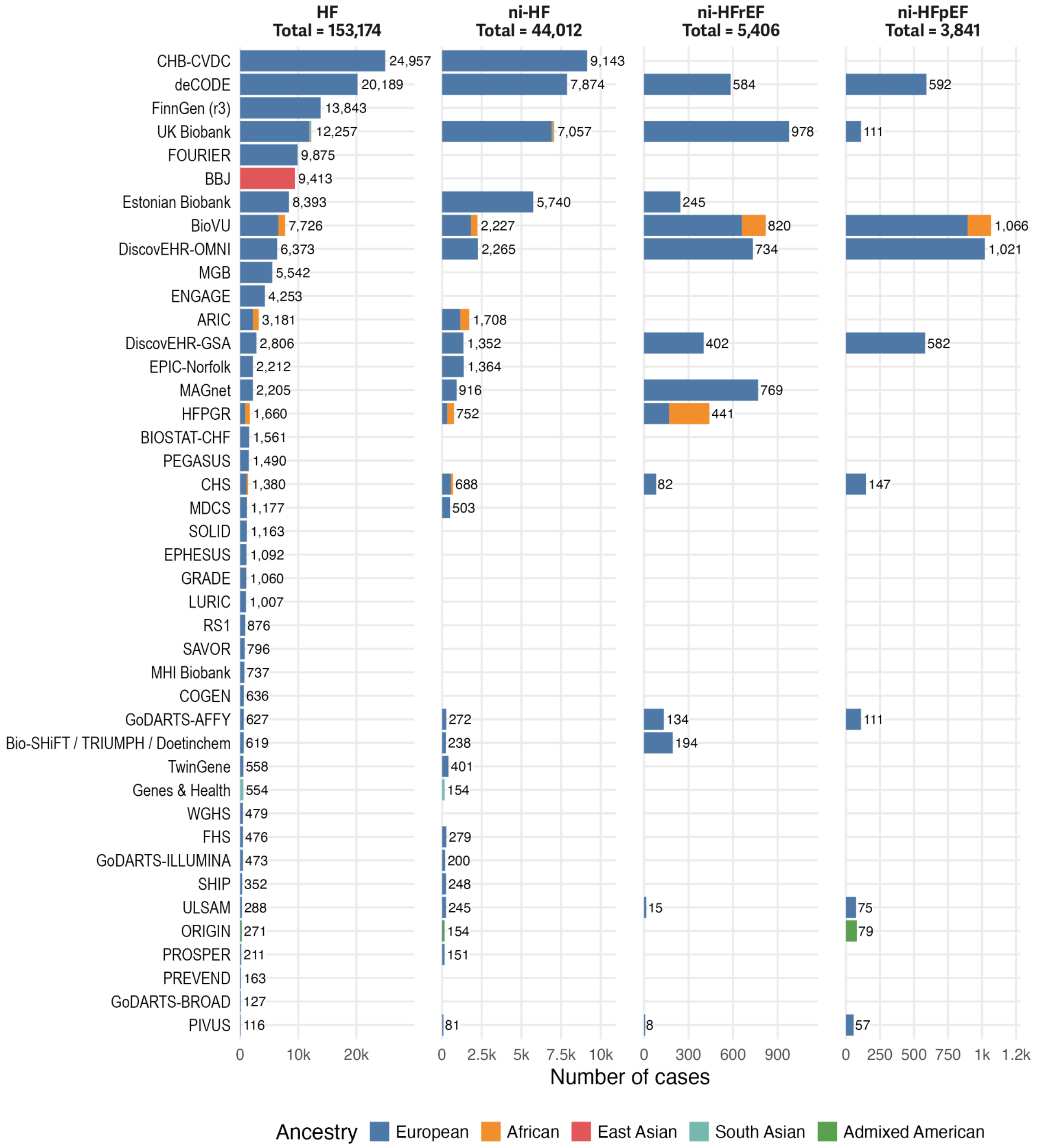


**Supplementary Figure 2.** Number of HF cases across cohorts participating in the present GWAS meta-analysis. HF: Heart failure, ni-HF: non-ischaemic HF, ni-HFrEF: non-ischaemic with reduced ejection fraction (LVEF <50%), ni-HFpEF: non-ischaemic with preserved ejection fraction (LVEF ≥50%).

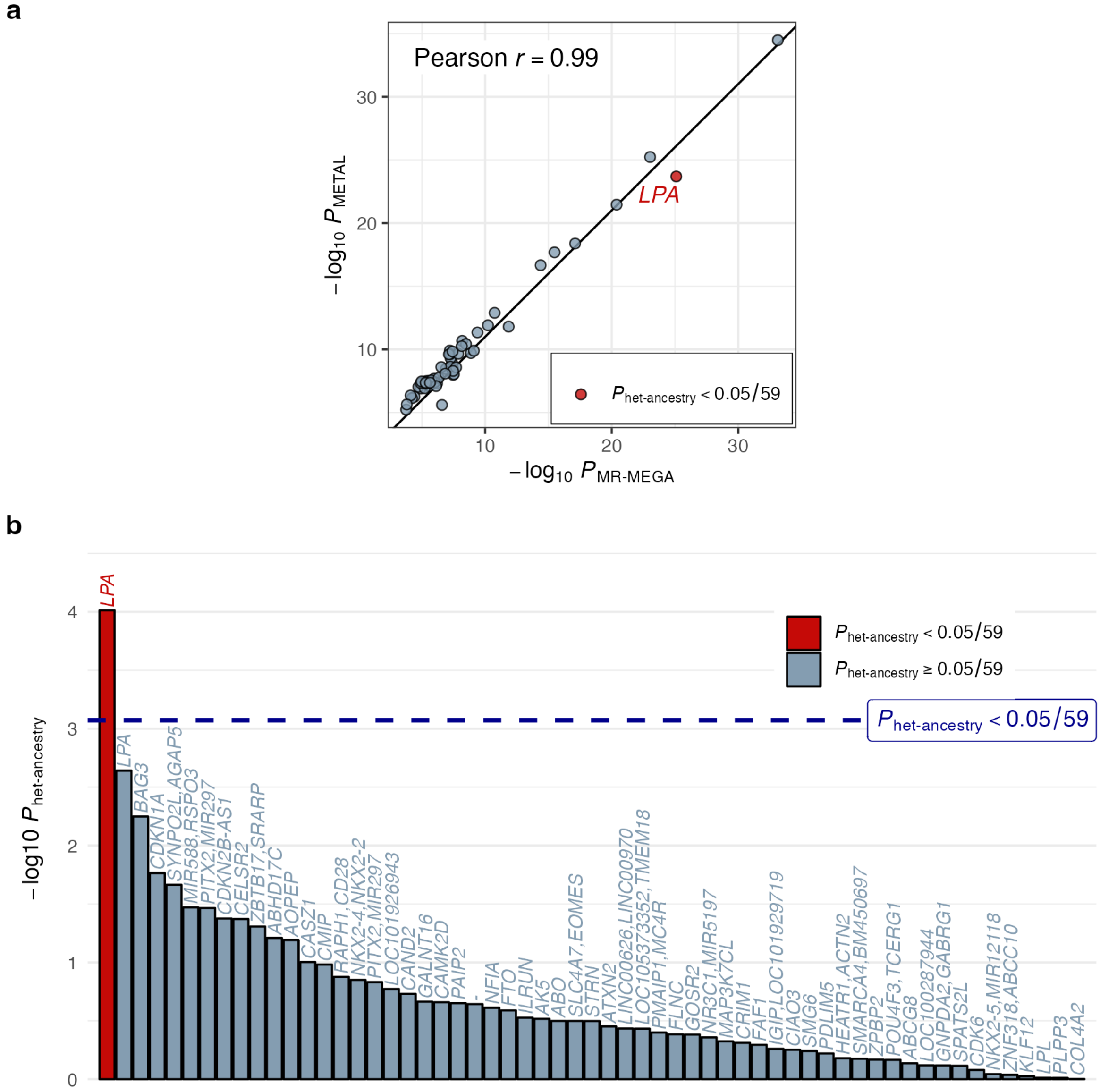


**Supplementary Figure 3. a.** *P* value for associations from multi-ancestry meta-analysis estimated with MR-MEGA and METAL for 59 conditionally independent lead variants associated with all-comer HF at $P < 5 \times 10^{-8}$. Lead variants with allelic effect heterogeneity surviving multiple testing correction ($P_{\text{het-ancestry}}$ < 0.05 / 59) are labelled with the nearest gene. **b.** *P* value for heterogeneity due to ancestry ($P_{het\text{-}ancestry}$) estimated using MR-MEGA for 59 conditionally independent lead variants associated with all-comer HF at $P < 5 \times 10^{-8}$. Variants are labelled with nearest gene(s), and colored based on $P_{het\text{-}ancestry}$ below / equal to or greater than Bonferroni-corrected alpha 0.05 / 59.

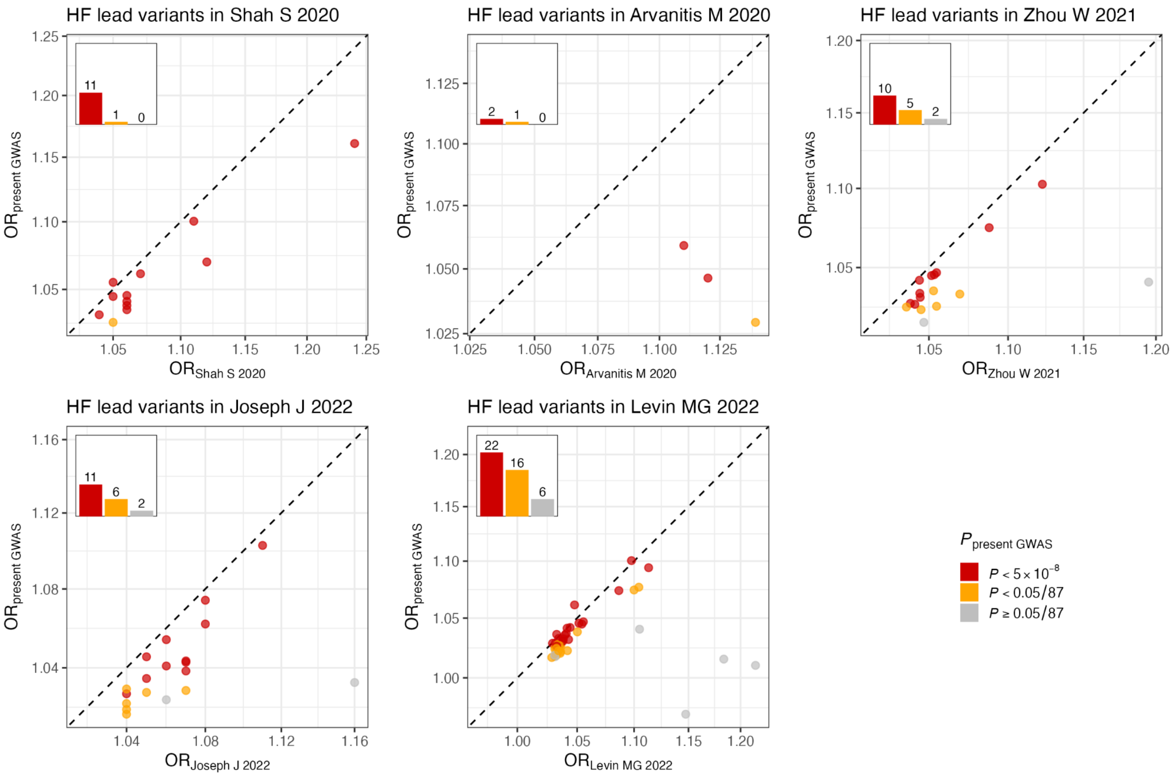


**Supplementary Figure 4.** Comparison of effect estimates of sentinel variants reported in previous heart failure GWAS (horizontal axis) by Shah S, Henry A, et al (2020)(Shah et al. 2020), Arvanitis M et al (2020)(Arvanitis et al. 2020), Zhou W et al (2021)(Zhou et al. 2022), Joseph J et al (2022), and Levin MG et al (2022)(Levin et al. 2022) with effect estimates from the present GWAS of HF (vertical axis).

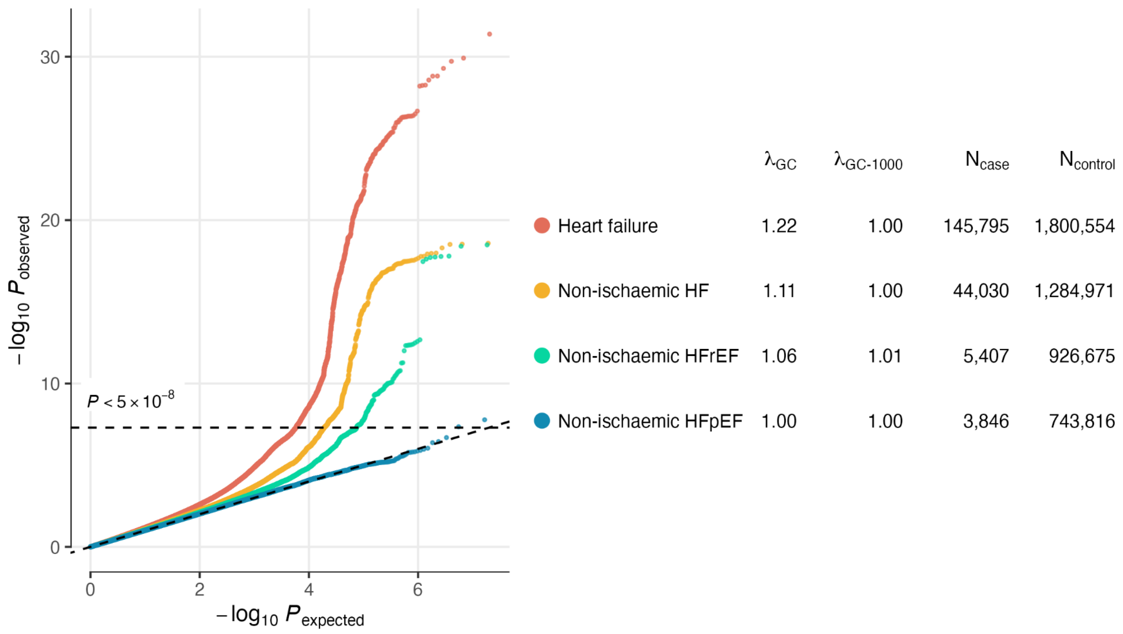


**Supplementary Figure 5.** Quantile-quantile plots of the GWAS meta-analysis results. $\lambda_{GC}$, genomic control statistics; $\lambda_{GC-1000}$, genomic control statistics assuming 1000 samples; $N_{case/control}$, Number of cases / controls.

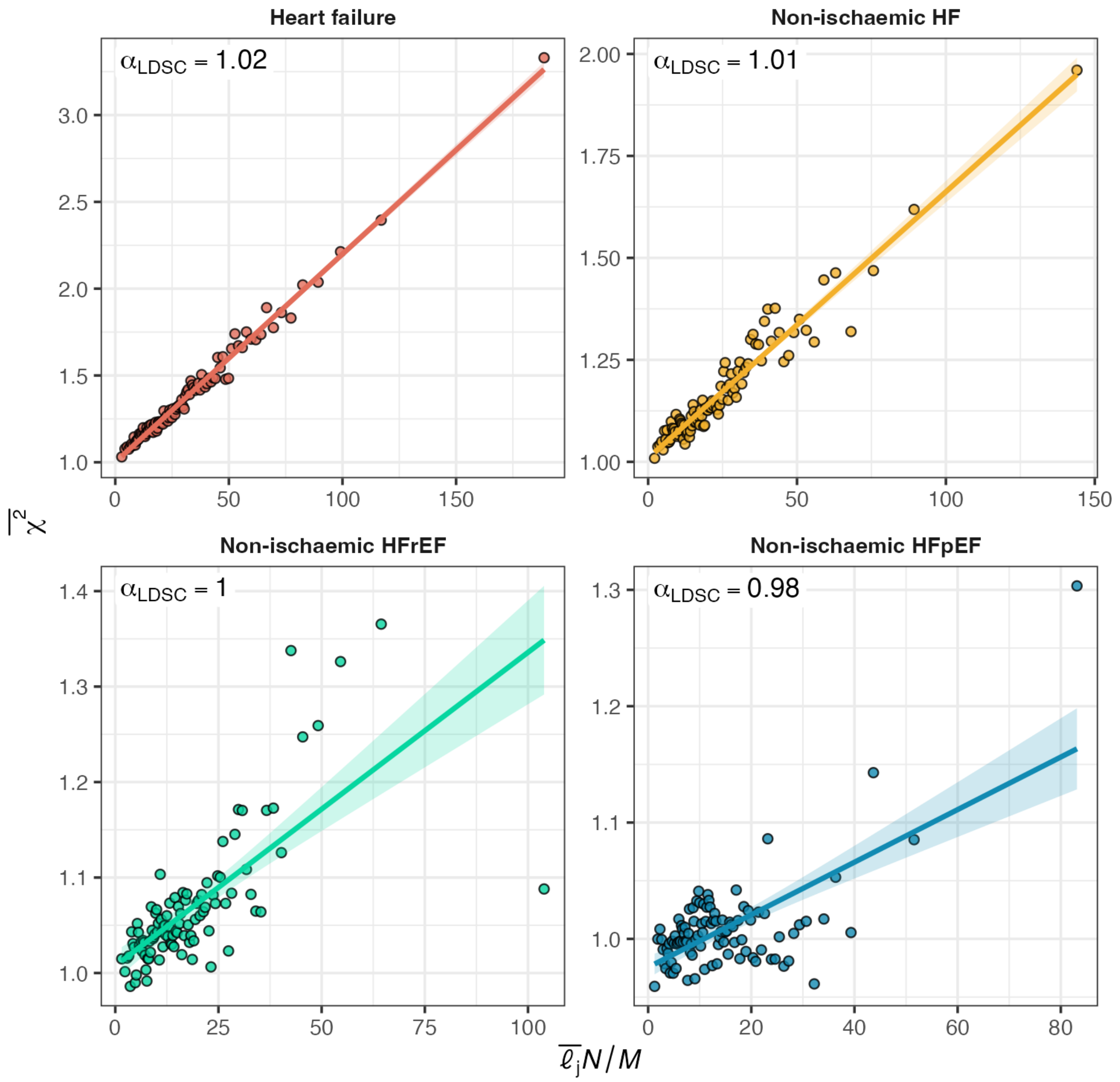


**Supplementary Figure 6.** Linkage disequilibrium score (LDSC) regression plots for percentiles of HapMap3 SNPs in the European 1000G reference panel. Each data point represents one percentile group in a given phenotype, with Y coordinate representing mean chi-squared statistics ($\overline{\chi^2}$) from the present GWAS meta-analysis in European ancestry subset, and X coordinate representing mean LD score ($\overline{\ell}_j$) times the number of GWAS samples (*N*) divided by the number of SNPs / markers (*M*) post-merging. The LDSC intercept ($\alpha_{LDSC}$) are annotated.

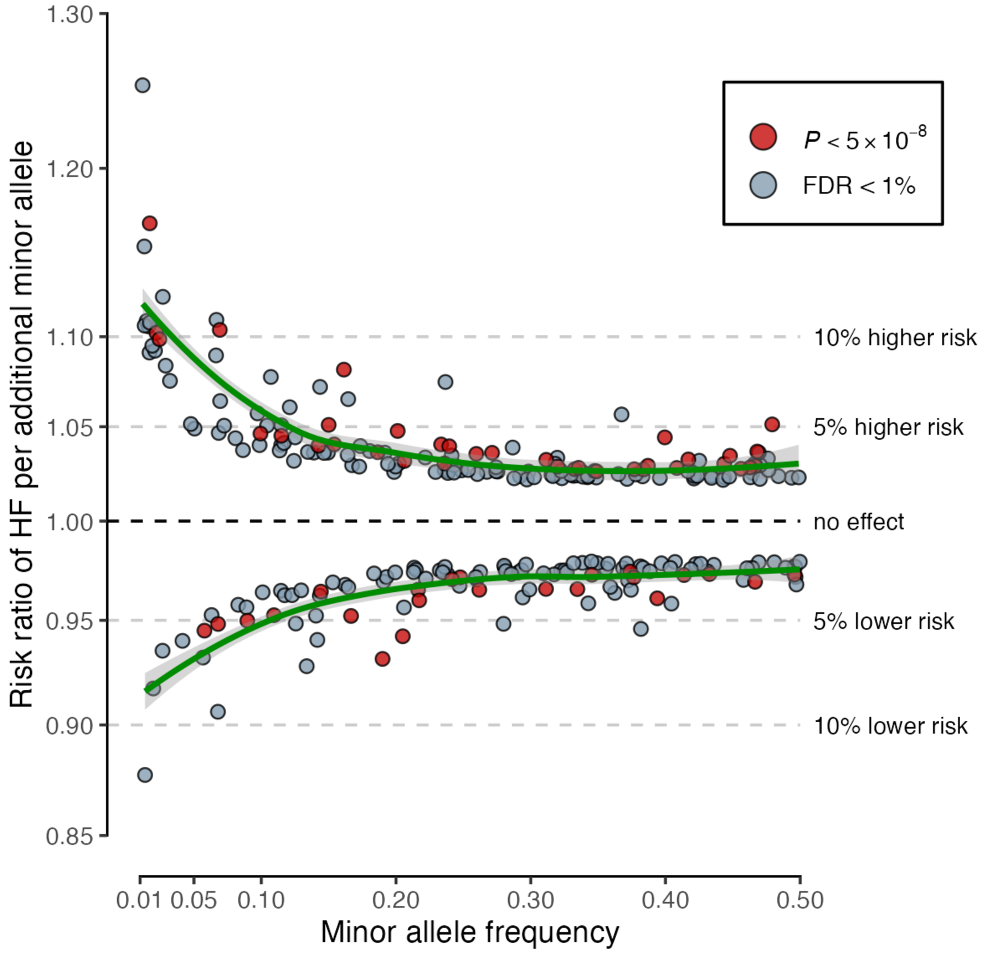


**Supplementary Figure 7.** Allelic architecture of conditionally independent variants for HF. Green lines represent local polynomial regression lines.

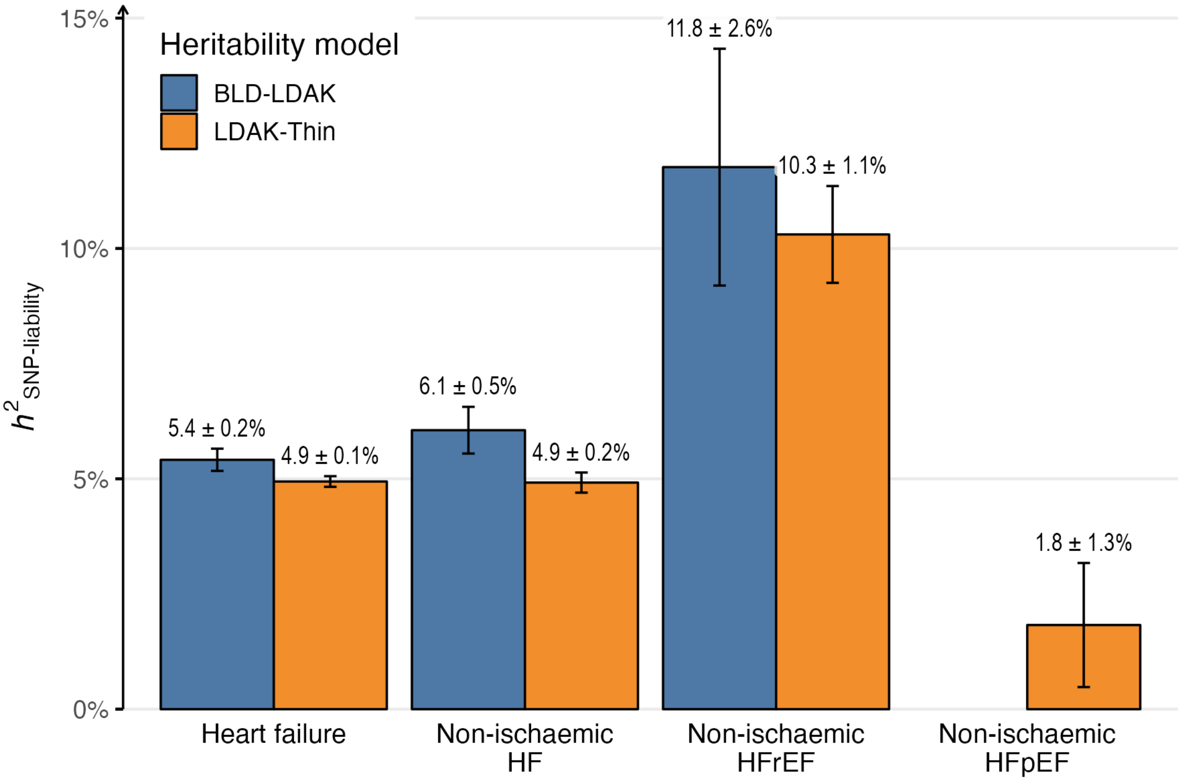


**Supplementary Figure 8.** SNP heritability on a liability scale ($h^2_{\text{SNP-liability}}$) estimates under 66-parameter BLD-LDAK and LDAK-Thin heritability models with European meta-analysis subset for heart failure phenotypes. The BLD-LDAK heritability estimate for non-ischaemic HFpEF was unavailable due to a limited sample size.

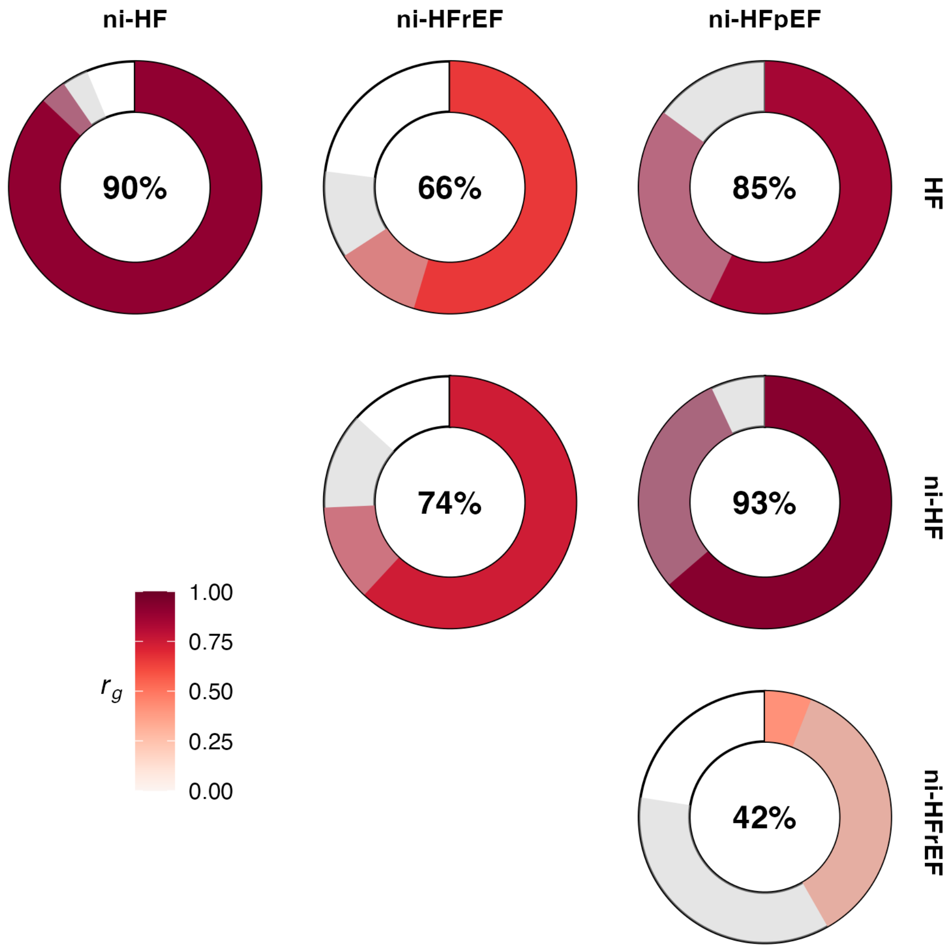


**Supplementary Figure 9.** Genetic correlation ($r_g$) between HF phenotypes. The red-shaded area represents point estimate (annotated as percentage in the centre of each circle). The grey shaded area represents 95% confidence intervals. For display, all estimates are constrained within 0 (no genetic correlation) to 1 (perfect generation correlation) range.

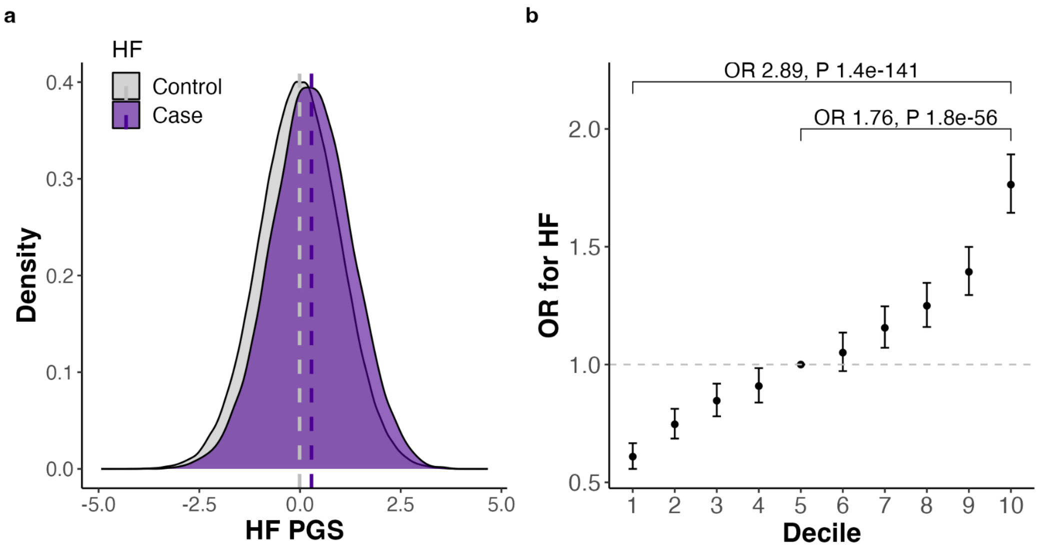


**Supplementary Figure 10.** a. Distribution of polygenic score for HF ($PGS_{HF}$) amongst individuals with (case) and without HF (control) in UK Biobank. b. Odds ratio for HF across deciles of $PGS_{HF}$ in UK Biobank.

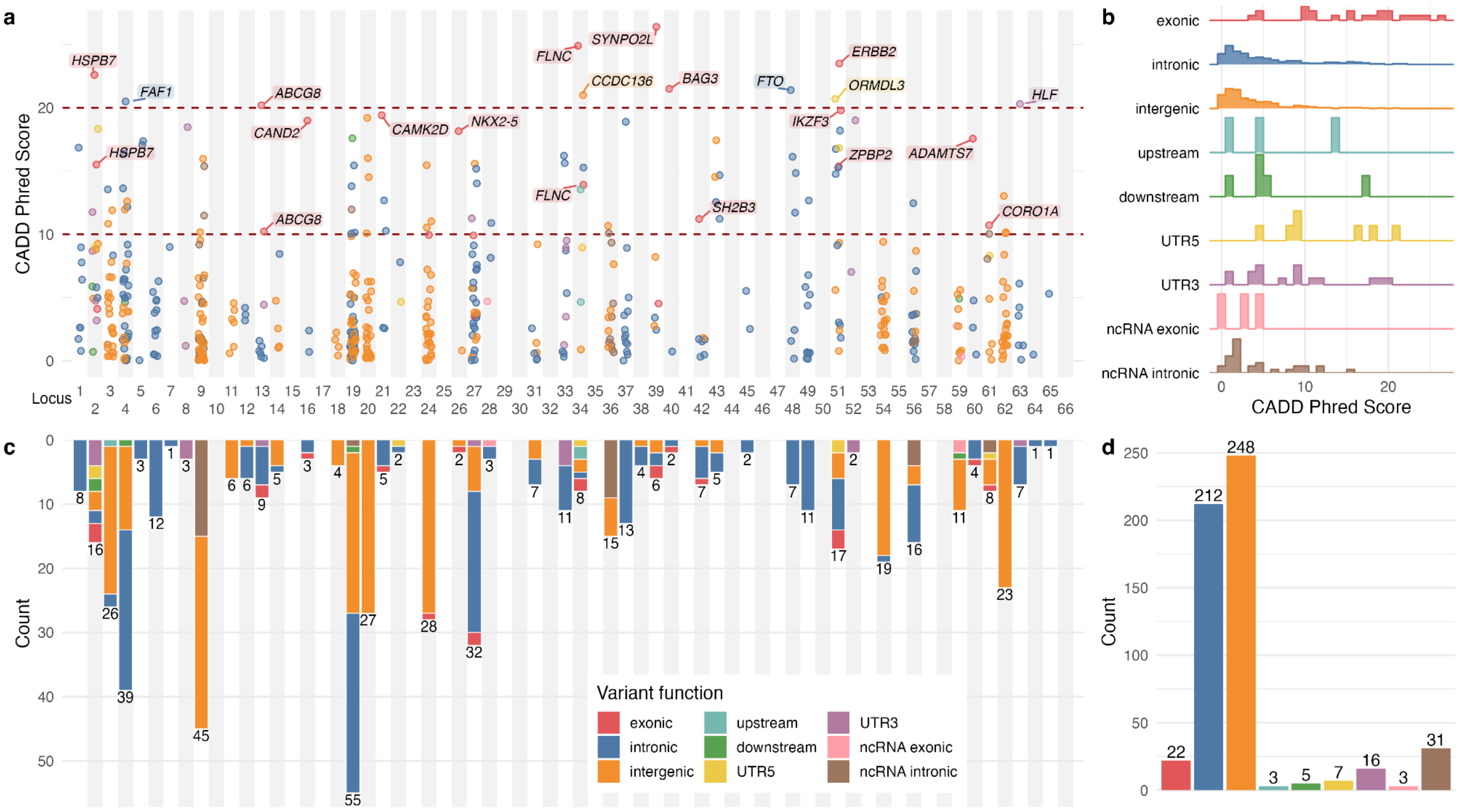


**Supplementary Figure 11. a.** Predicted deleteriousness of 547 finemapped variants within 95% credible sets across 66 independent genomic loci for HF as measured by Combined Annotation Dependent Depletion (CADD) Phred score. Nearest genes of finemapped variants with CADD Phred score >20 and finemapped exonic variants with CADD Phred score > 10 are labelled. **b.** Distribution of CADD Phred Score amongst fine-mapped variants stratified by function c. Locus-wide and d. genome-wide counts of fine-mapped variants stratified by variant function

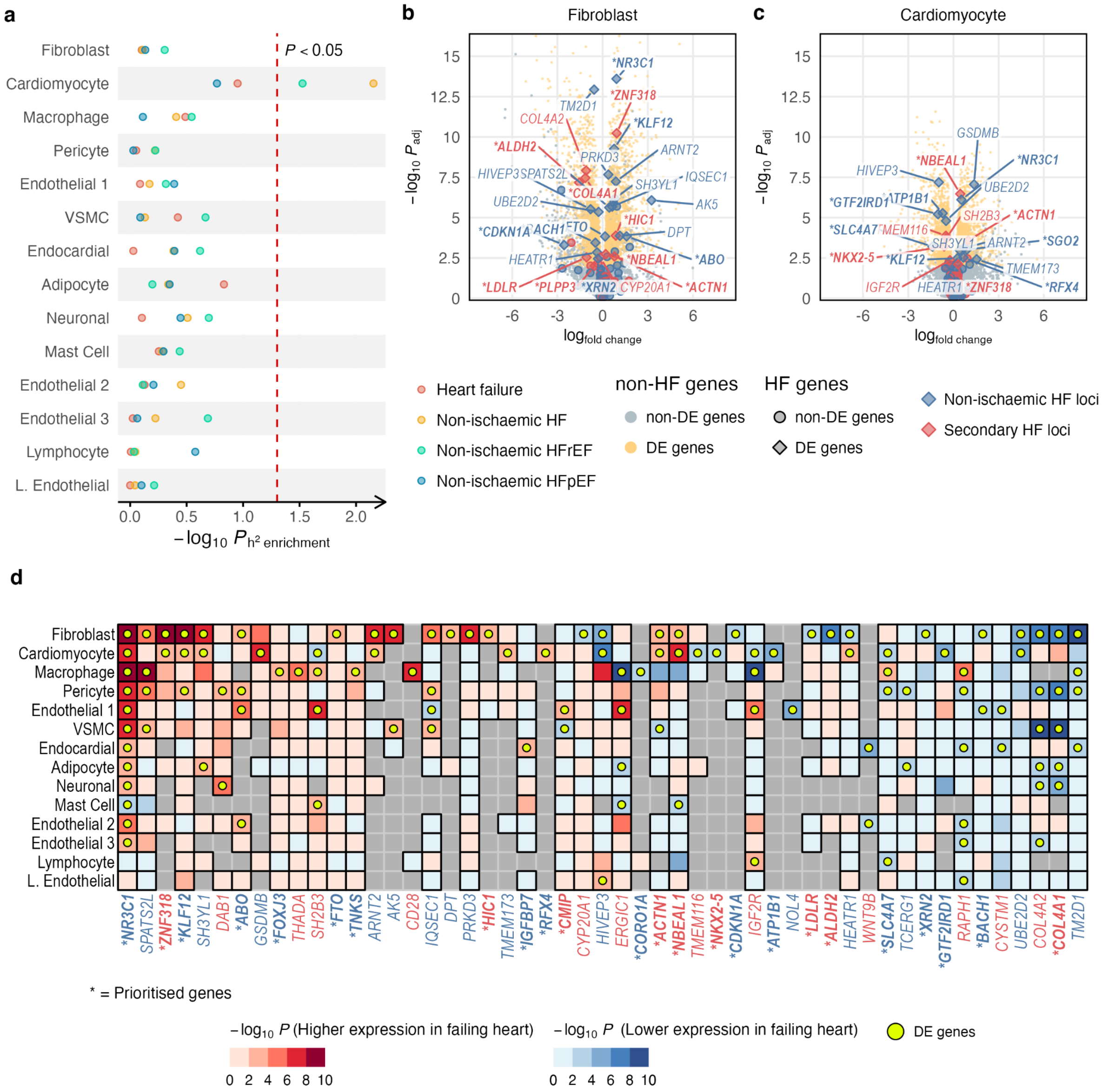


**Supplementary Figure 12. a**. Heritability enrichment across cell types in non-failing heart samples. **b-c.** Volcano plots showing differential gene expression between cell types (fibroblasts and cardiomyocytes) from failing and non-failing heart samples. Candidate and prioritised HF genes (the latter is prefixed with *) that are differentially expressed (DE) are highlighted. **d.** Cross-cell types differential gene expression profile for 51 candidate or prioritised HF genes that are differentially expressed in at least 1 cardiac cell type. Grey tiles = no data.

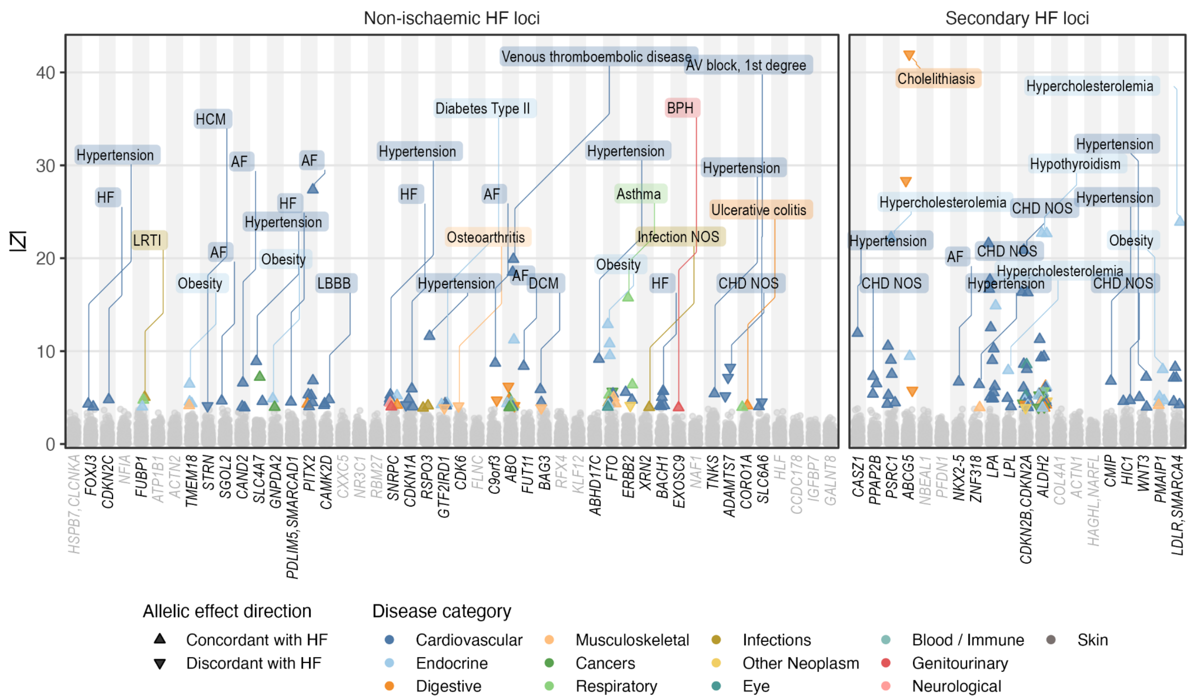


**Supplementary Figure 13.** Phenome-wide association estimates in absolute Z-score (vertical axis) between 294 diseases and lead variants across 66 HF loci (horizontal axis). Estimates which survived multiple-testing correction at FDR < 1% are highlighted, with top associated phenotype (largest absolute Z score) per locus labelled. Loci without any FDR-passing association are greyed out.

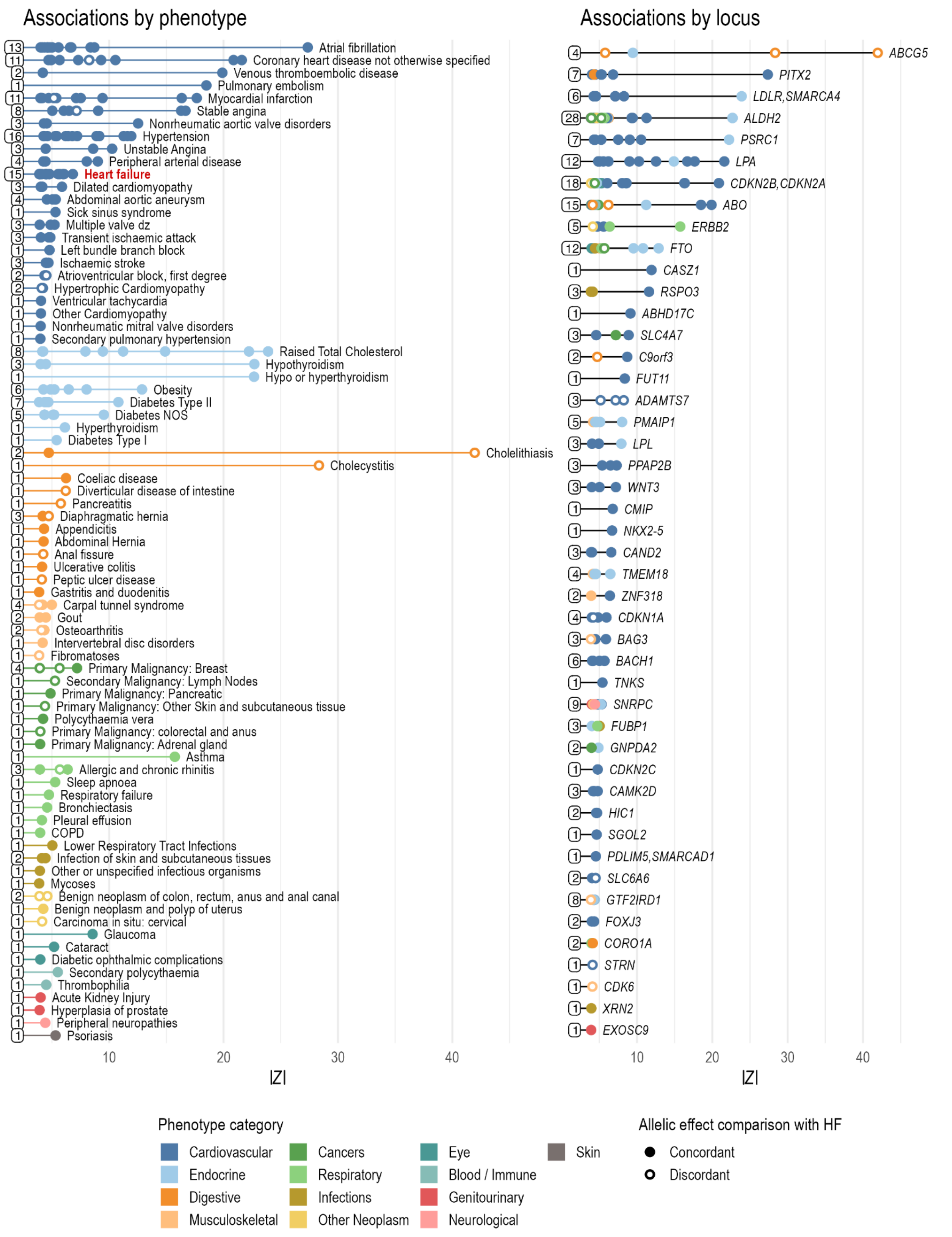


**Supplementary Figure 14.** Counts of associations between 79 (out of 294) disease phenotypes and 46 (out of 66) HF loci which survived the multiple testing adjustment at FDR <1%, stratified by phenotype and locus. The position of each bullet points along the X-axis represents strength of association as measured by absolute *Z* scores (|Z|). The base of the connecting line is labelled by the total number of associations for a given phenotype or locus.

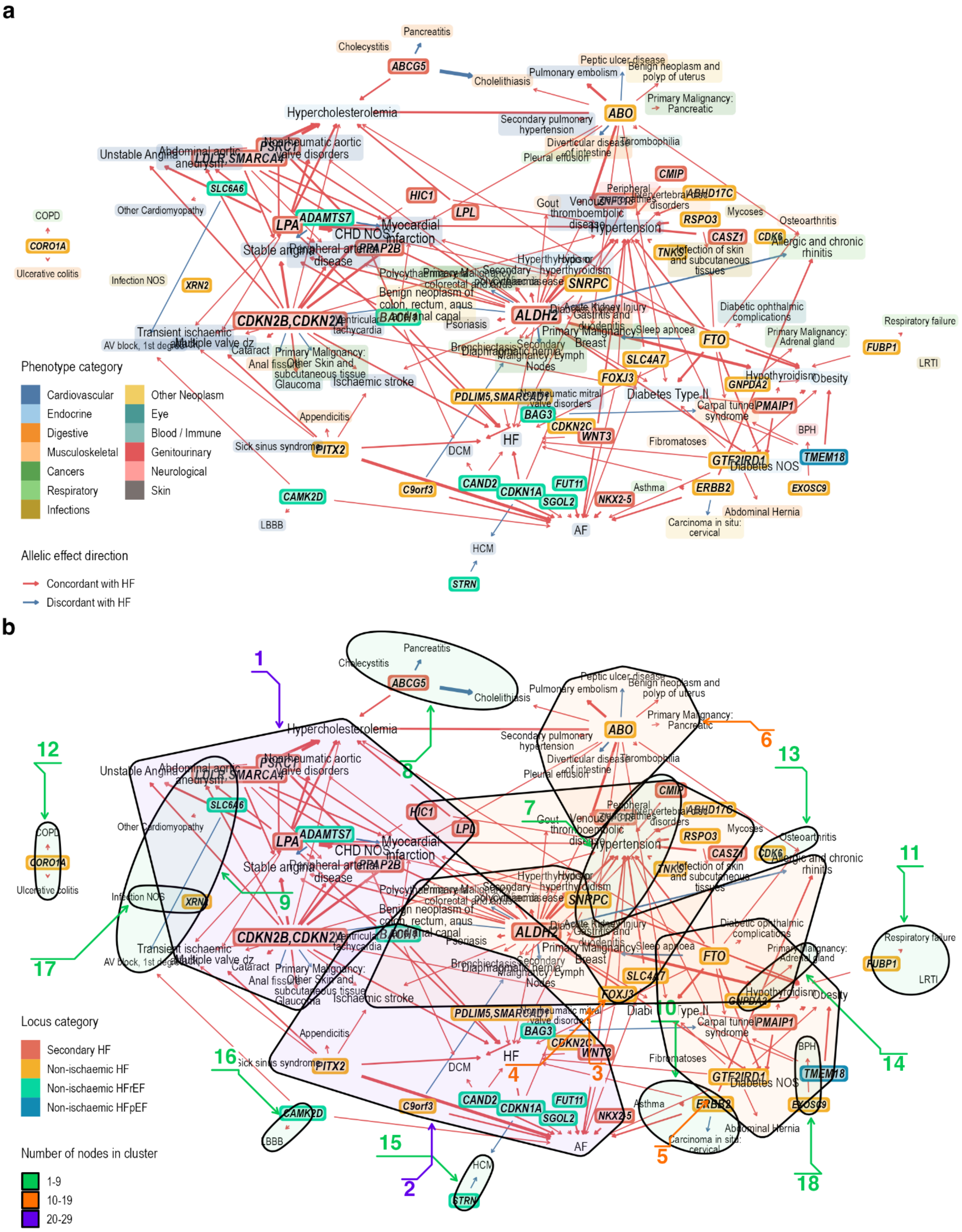


**Supplementary Figure 15. a.** Pleiotropy network from the 207 genotype-phenotype associations (edges) across 79 unique diseases (black text) and 46 HF locus (white text with black background) which passed FDR <1% correction from the PheWAS analysis. **b.** 18 node clusters formed from the pleiotropy network identified through the walktrap algorithm. For a full description of how the network was constructed, see **Methods**.

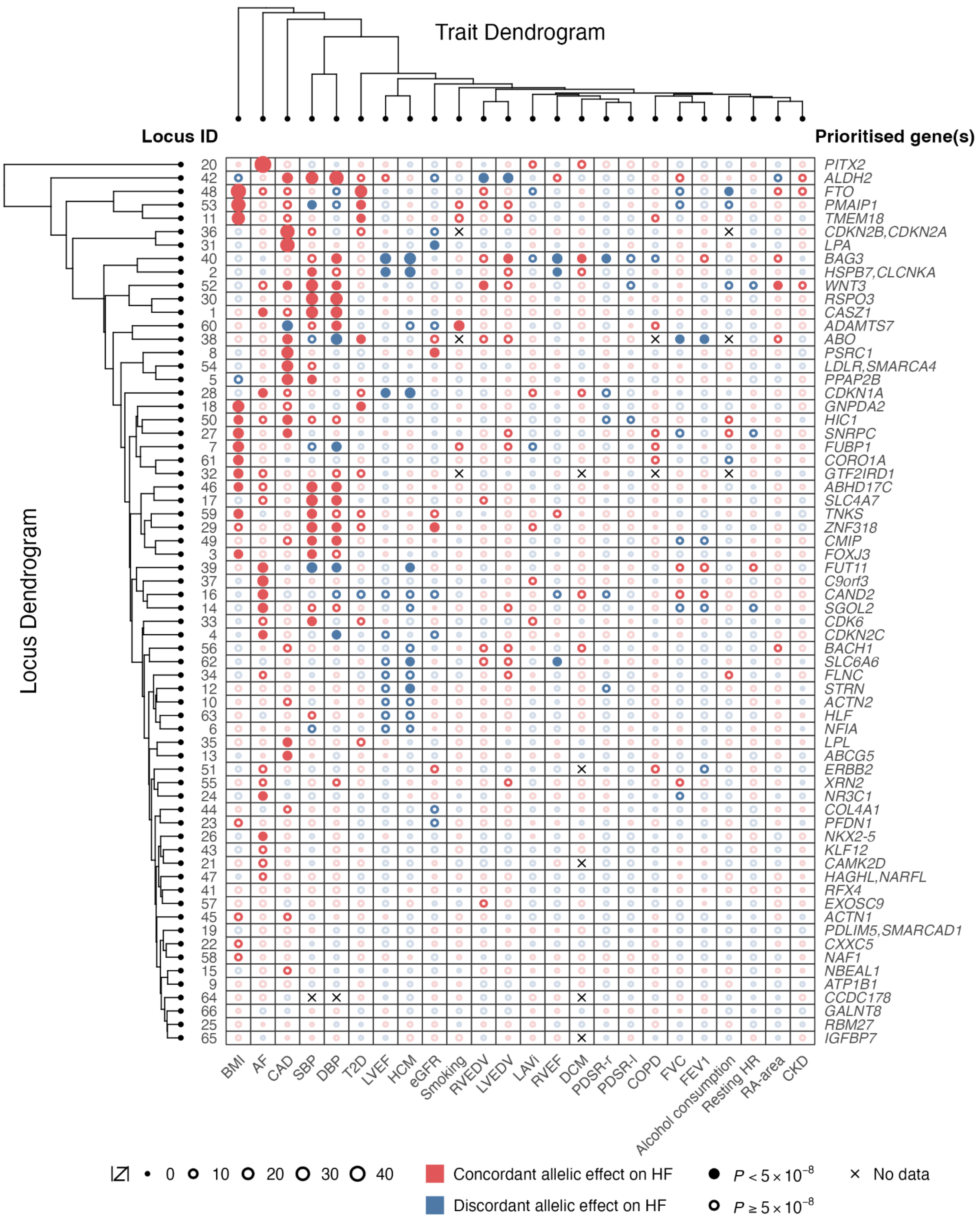


**Supplementary Figure 16.** Pleiotropic effects of lead variants in 66 HF loci (rows) across 24 GWAS traits (columns). Loci and traits were arranged based on hierarchical agglomerative clustering results, represented by dendrograms on the edges of the plot. Non-transparent circles represent associations which survived multiple testing adjustments at FDR < 1%.

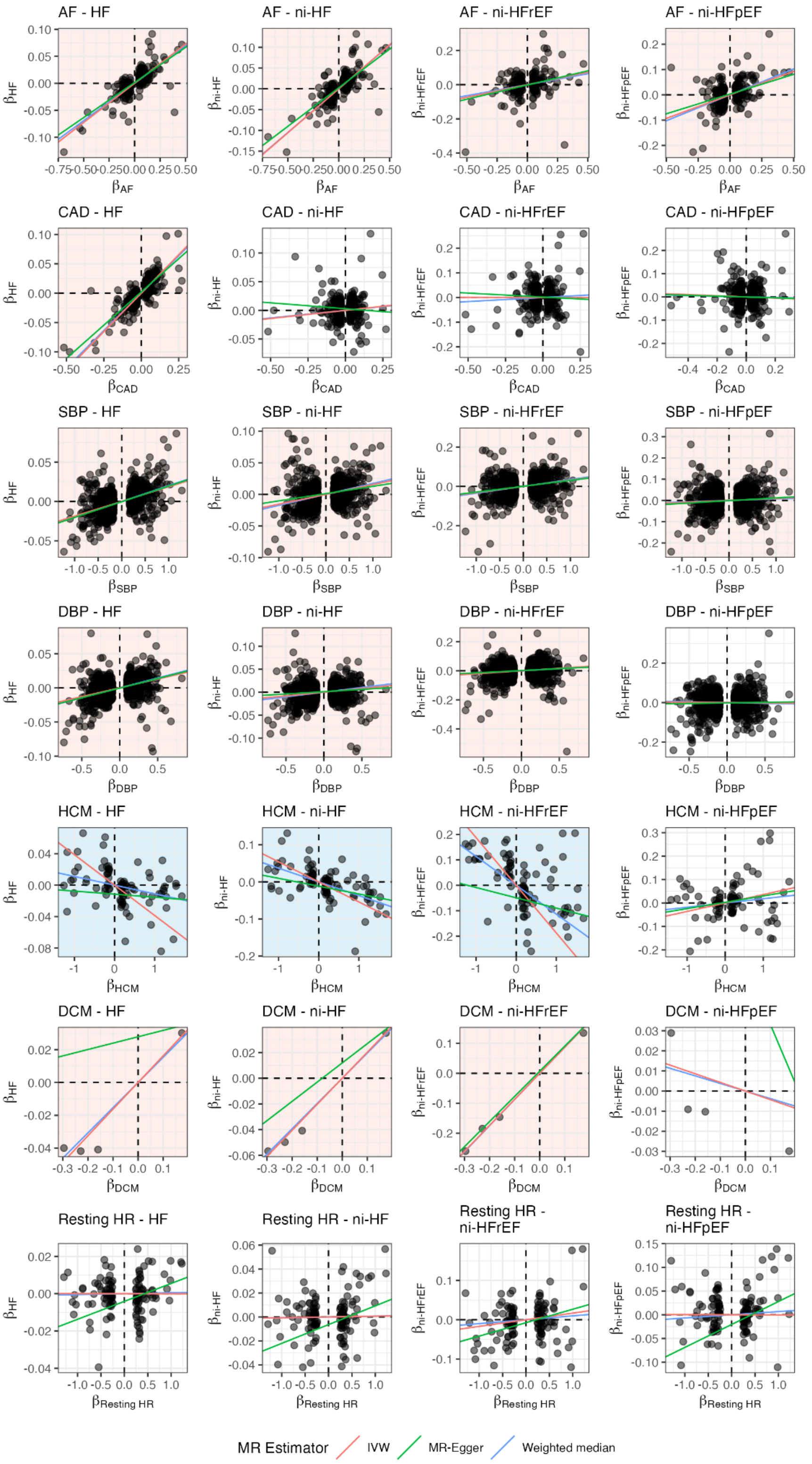

**Supplementary Figure 17.** Cross plot of estimated effect estimates of genetic instruments used in Mendelian randomisation (MR) analysis on exposure (horizontal axis) and on outcome traits (vertical axis). MR estimates passing false discovery rate (FDR) <1% as estimated using the inverse-variance weighted (IVW) estimator with consistent direction of effect estimated using MR-Egger and weighted median estimators are highlighted with light blue (risk-reducing effect) or light red (risk-increasing effect) background. The figure shows the estimated effect of selected cardiovascular and cardiomyopathies exposure traits. Trait abbreviations are provided in **Supplementary Table 23**.

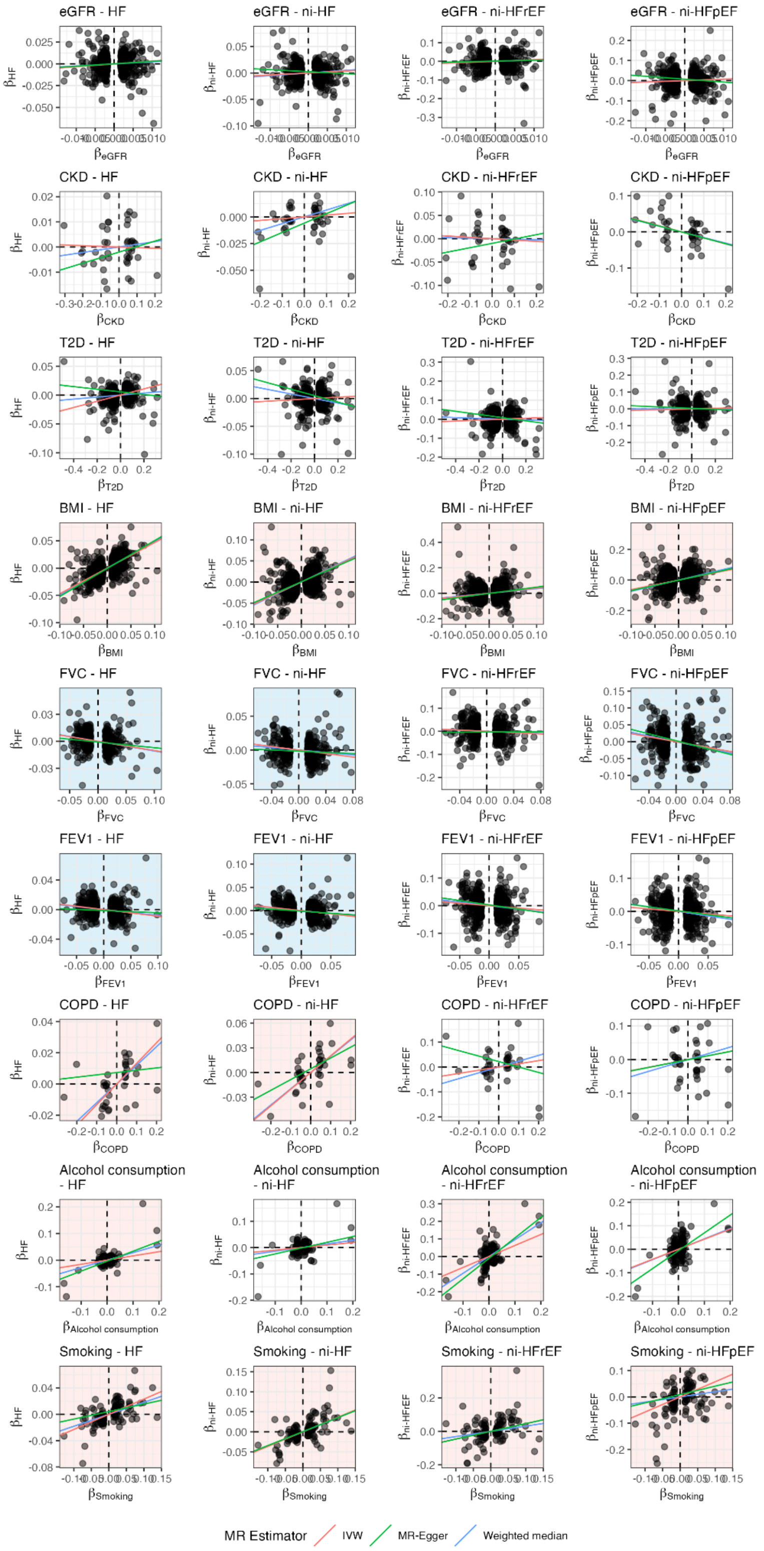


**Supplementary Figure 18.** Cross plot of estimated effect estimates of genetic instruments used in Mendelian randomisation (MR) analysis on exposure (horizontal axis) and on outcome traits (vertical axis). MR estimates passing false discovery rate (FDR) <1% as estimated using the inverse-variance weighted (IVW) estimator with consistent direction of effect estimated using MR-Egger and weighted median estimators are highlighted with light blue (risk-reducing effect) or light red (risk-increasing effect) background. The figure shows the estimated effect of selected metabolic, renal, respiratory and behavioural exposure traits. Trait abbreviations are provided in **Supplementary Table 23**.

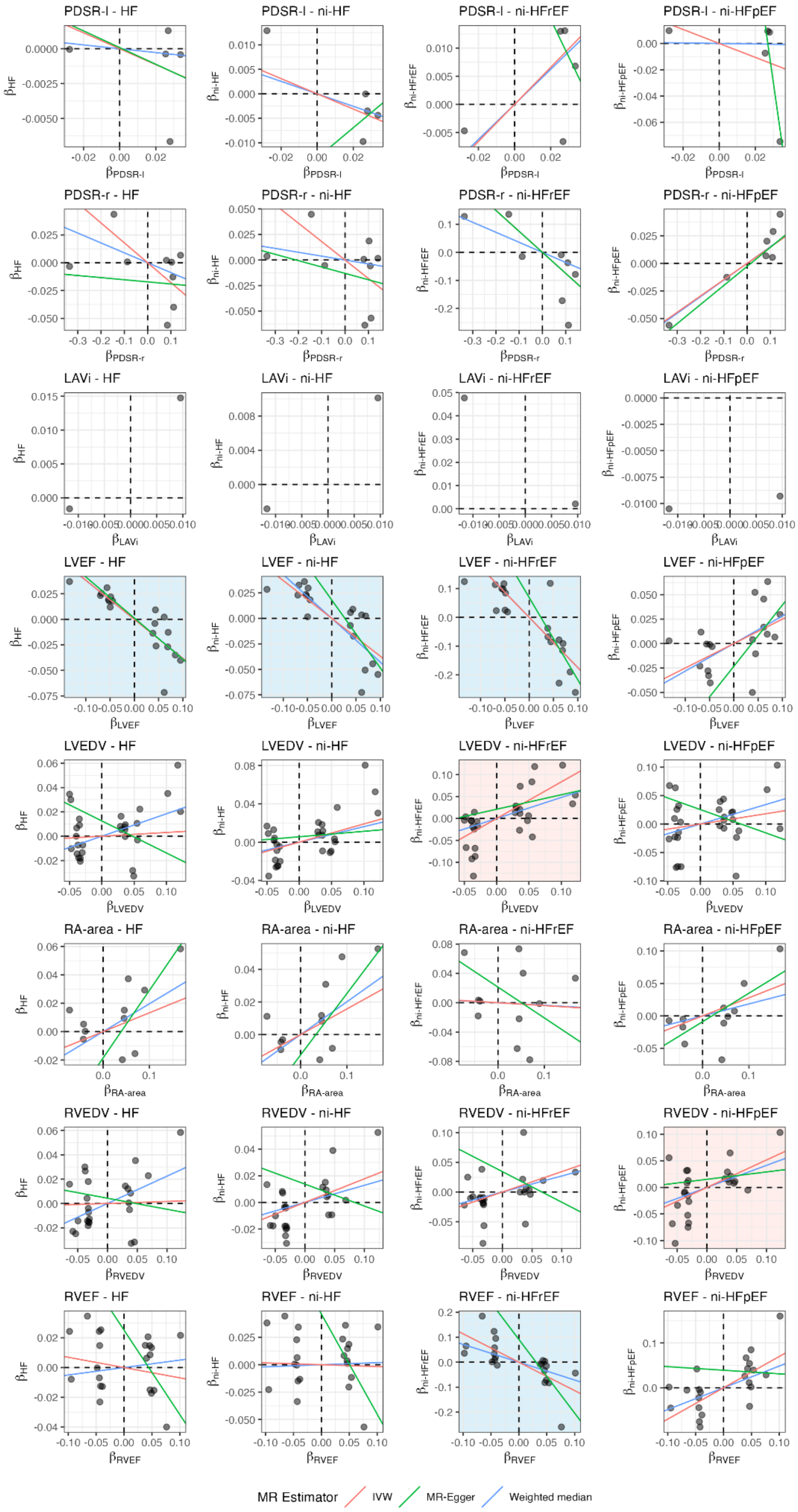


**Supplementary Figure 19.** Cross plot of estimated effect estimates of genetic instruments used in Mendelian randomisation (MR) analysis on exposure (horizontal axis) and on outcome traits (vertical axis). MR estimates passing false discovery rate (FDR) <1% as estimated using the inverse-variance weighted (IVW) estimator with consistent direction of effect estimated using MR-Egger and weighted median estimators are highlighted with light blue (risk-reducing effect) or light red (risk-increasing effect) background. The figure shows the estimated effect of selected cardiac functions derived from cardiac magnetic resonance imaging. Trait abbreviations are provided in **Supplementary Table 23**.

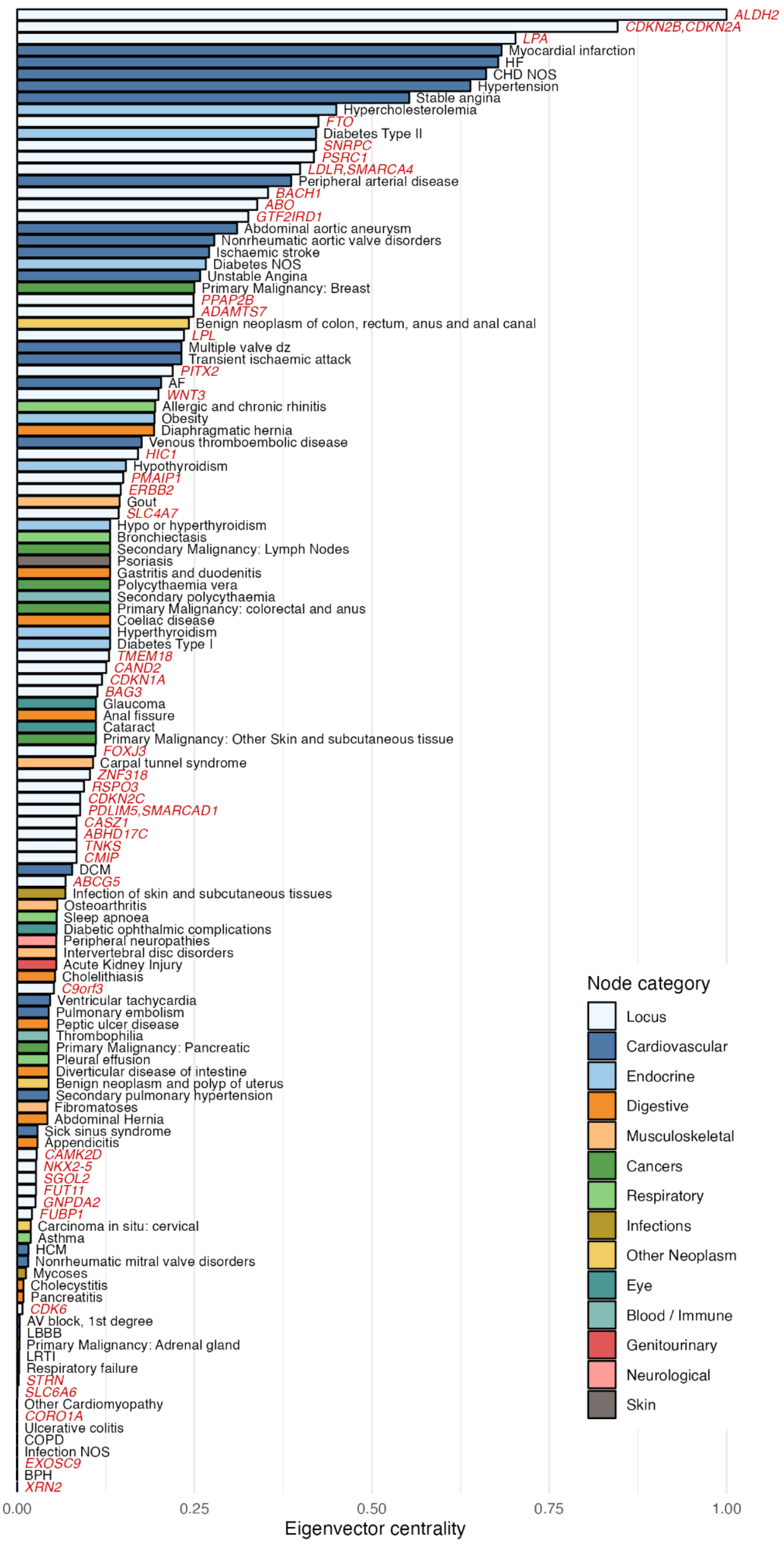


**Supplementary Figure 20.** Eigenvector centrality score of 46 unique loci (labelled by mapped genes in red colour) and 79 unique diseases from the PheWAS FDR <1% set that were included in the pleiotropy network analysis.
