## Supplementary Note for "Mapping the aetiological foundations of the heart failure spectrum using human genetics"

#### Phenotype Definition

To define heart failure (HF) phenotypes, we developed a multi-modal, rule-based phenotyping algorithm using a combination of physician adjudication, codes for diagnosis or procedure in electronic health record (EHR), and left ventricular ejection fraction (LVEF) measure. The algorithm is divided into two steps as described below.

##### Step 1: Define phenotype classifiers

For each study participant, define status (*TRUE / FALSE*) for the following phenotype classifiers:

A. All-cause heart failure (HF)

| Preference -ranked options | Manual adjudication | Diagnosis / procedure codes* | Medication† | Free text‡ | Imaging | Rule |
|---|---|---|---|---|---|---|
| Option 1 | YES | NO | NO | NO | N/A | Manually adjudicated HF diagnosis |
| Option 2 | NO | YES | YES | YES | N/A | ≥3 instances of diagnosis codes* AND HF medication† AND free text |
| Option 3 | NO | YES | YES | NO | N/A | ≥1 instances of diagnosis codes AND HF medication |
| Option 4 | NO | YES | NO | NO | N/A | ≥3 instances of diagnosis codes |

* see **Appendix 1** ICD-9 and ICD-10 code lists
† see **Appendix 2** for list of HF medications
‡ see **Appendix 3** for list of free text search strings

B. HF following coronary artery disease, valvular or congenital heart disease (Secondary HF)

| Preference ranked options | Manual adjudication* | Diagnosis / procedure codes† | Medication | Free text | Imaging | Rule |
|---|---|---|---|---|---|---|
| Option 1 | YES | NO | N/A | N/A | N/A | Manually adjudicated diagnosis* |
| Option 2 | NO | YES | N/A | N/A | N/A | ≥1 instances of ICD codes† |

* for manual adjudication, the relevant diagnoses are as follows:

*Coronary artery disease*
If possible, please apply a hard definition of coronary artery disease defined as:

- Myocardial infarction
- Coronary artery revascularization (PCI or CABG) procedure

Otherwise please use study level definitions to define coronary artery disease status.

*Coronary artery disease, valvular or congenital heart disease*
Please define as the presence of the following diagnoses:

- Severe primary valve disease defined by a relevant surgical or percutaneous valve procedure
- Any congenital malformation of the heart or great vessels defined by a diagnosis, or relevant surgical or percutaneous procedure
- Any rheumatic heart valve disease defined by diagnosis

† see **Appendix 4** for ICD-9, ICD-10, OPCS4 and CPT code lists

Note: This list contains

- diagnosis of congenital malformation of the heart or great vessels
- diagnosis of rheumatic heart valve disease
- diagnosis of myocardial infarction
- surgical or percutaneous procedure for coronary artery revascularization
- surgical or percutaneous procedure for valvular heart disease
- surgical or percutaneous procedure for congenital malformation of the heart or great vessels

C. History of left ventricular systolic dysfunction (Ever LVSD)

| **Preference ranked options** | **Manual adjudicat ion*** | **Diagnosis / procedure codes†** | **Medicat ion** | **Free text** | **Imaging** | **Rule** |
|---|---|---|---|---|---|---|
| Option 1 | NO | NO | N/A | N/A | YES | LVEF measure <50% |
| Option 2 | YES | NO | N/A | N/A | NO | Manually adjudicated diagnosis* |
| Option 3 | NO | YES | N/A | N/A | NO | ≥1 instances of ICD codes |

* for manual adjudication, the relevant diagnoses are as follows:

- Heart failure with reduced ejection fraction
- Heart failure with mid-range ejection
- Systolic heart failure
- Dilated cardiomyopathy
- Peripartum cardiomyopathy
- Alcohol cardiomyopathy
- Inflammatory cardiomyopathy
- Chemotherapy-related cardiotoxic

† see **Appendix 5** for ICD-9, ICD-10, OPCS4 and CPT code lists
‡ Positive predictive value (PPV) and sensitivity are estimated with clinically-adjudicated case status and imaging data from the BioVU study

D. Left ventricular ejection fraction ≥ 50% (LVEF ≥50%)

| Manual adjudication | Diagnosis codes | Medication | Free text | Imaging | Preference ranked options | Rule |
|---|---|---|---|---|---|---|
| N/A | N/A | N/A | N/A | YES | Option 1 | LVEF measure ≥50%* |

*LVEF measure ≥50% on cardiac imaging (any modality) *at or after the time of first HF diagnosis or evidence of HF in the record.*

#### Step 2: Define target phenotypes

Using a combination of boolean states from the four phenotype classifiers above, the step 2 of the phenotyping algorithm then classifies each study participant as a *Case*, *Control*, or to be *Excluded* from analysis for each target phenotype as outlined in **Appendix 6** and **Appendix 7**. The exclusion criteria in the phenotyping algorithm were designed to improve discovery in a case control association analysis by removing potential competing risk factors or aetiologies from the analysis.

#### HERMES Consortium Contributors

The following names are members of the HERMES Consortium at the time of the analysis, listed in alphabetical order by surname:

Erik Abner, Lance Adams, Peter Almgren, Charlotte Andersson, Krishna G. Aragam, Johan Ärnlöv, Folkert W. Asselbergs, Geraldine Asselin, Joshua D. Backman, John Baksi, Paul J.R. Barton, Traci M. Bartz, Traci Bartz, Kiran J. Biddinger, Mary L. Biggs, Heather L. Bloom, Eric Boersma, Isabelle Bond, Maxime Bos, Jeffrey Brandimarto, Michael R. Brown, Hans-Peter Brunner-La Rocca, Rachel Buchan, Leonard Buckbinder, Douglas Cannie, Thomas P. Cappola, David J. Carey, Mark D. Chaffin, Philippe Charron, Daniel I. Chasman, Xing Chen, Sandesh Chopade, William Chutkow, John G.F. Cleland, James P. Cook, Stuart A. Cook, Tomasz Czuba, Trégouët David-Alexandre, Simon Dedenus, Abbas Dehghan, Graciela E. Delgado, Spiros Denaxas, Alexander S. Doney, Marie-Pierre Dubé, J. Gustav Smith, Samuel C. Dudley, Michael E. Dunn, Marcus Dörr, Patrick T. Ellinor, Perry Elliott, Gunnar Engström, Villard Eric, Tõnu Esko, Eric H. Farber-Eger, Eric Farber-Eger, Stephan B. Felix, Chris Finan, Sarah Finer, Ian Ford, René Fouodjio, Catherine Francis, Sophie Garnier, Mohsen Ghanbari, Sahar Ghasemi, Jonas Ghouse, Vilmantas Giedraitis, Franco Giulianini, John S. Gottdiener, Jasmine Gratton, Stefan Gross, Hongsheng Gui, Karl Guo, Rebecca Gutmann, Daníel F. Guðbjartsson, Christopher M. Haggerty, Brian Halliday, Åsa K. Hedman, Anna Helgadottir, Harry Hemingway, Albert Henry, Hans Hillege, Aroon D. Hingorani, Hilma Holm, Craig L. Hyde, Sara Hägg, Hanane Issa, Jaison Jacob, Deleuze Jean-François, J. Wouter Jukema, Frederick Kamanu, Isabella Kardys, Ravi Karra, Maryam Kavousi, Kay-Tee Khaw, Jorge R. Kizer, Jorge Kizer, Marcus E. Kleber, Andrea Koekemoer, Bill Kraus, Karoline Kuchenbaecker, Lars Køber, Yi-Pin Lai, David Lanfear, Chim C. Lang, Claudia Langenberg, Michael Lee, Honghuang Lin, Lars Lind, Cecilia M. Lindgren, Peter P. Liu, Barry London, Brandon D. Lowery, Brandon Lowery, Jian'an Luan, Steven A. Lubitz, R. Thomas Lumbers, Patrik Magnusson, Anubha Mahajan, Anders Malarstig, Douglass Mann, Kenneth B. Margulies, Nicholas A. Marston, Nicholas Marston, Hillary Martin, Kathryn McGurk, John J.V. McMurray, Olle Melander, Giorgio Melloni, Andres Metspalu, David Miller, Xiaodong Mo, Ify R. Mordi, Michael P. Morley, Andrew P. Morris, Andrew D. Morris, Alanna C. Morrison, Lori Morton, Winfried März, Michael W. Nagle, Christopher P. Nelson, Alexander Niessner, Teemu Niiranen, Raymond Noordam, Michela Noseda, Christoph Nowak, Michelle L. O'Donoghue, Declan P O'Regan, Anjali T. Owens, Colin N. A. Palmer, Antonis Pantazis, Helen M. Parry, Guillaume Paré, Markus Perola, Louis-Philippe Lemieux Perreault, Charron Philippe, Marie Pigeyre, Eliana Portilla-Fernandez, Sanjay K. Prasad, Bruce M. Psaty, Kenneth M. Rice, Paul M. Ridker, Simon P. R. Romaine, Carolina Roselli, Jerome I. Rotter, Christian T. Ruff, Mark S. Sabatine, Mark Sabatine, Neneh Sallah, Perttu Salo, Veikko Salomaa, Nilesh J. Samani, Naveed Sattar, A. Floriaan Schmidt, Jessica van Setten, Sonia Shah, Svati Shah, Alaa A. Shalaby, Akshay Shekhar, Diane T. Smelser, Nicholas L. Smith, J. Gustav Smith, Garnier Sophie, Doug Speed, Sundararajan Srinivasan, Kari Stefansson, Steen Stender, David J. Stott, Garðar Sveinbjörnsson, Per Svensson, Daniel I. Swerdlow, Petros Syrris, Mari-Liis Tammesoo, Jean-Claude Tardif, Upasana Tayal, Maris Teder-Laving, Alexander Teumer, Pantazis I Theotokis, Guðmundur Thorgeirsson, Unnur Thorsteinsdottir, Christian Torp-Pedersen, Vinicius Tragante, Stella Trompet, Andre G. Uitterlinden, Ramachandran S. Vasan, Felix Vaura, Abirami Veluchamy, Monique Verschuuren, Niek Verweij, Adriaan A. Voors, Marion van Vugt, Uwe Völker, Lars Wallentin, Yunzhang Wang, James S. Ware, James Ware, Nicholas J. Wareham, Dawn Waterworth, Peter E. Weeke, Peter Weeke, Raul Weiss, Quinn Wells, Harvey White, Kerri L. Wiggins, Jemma B. Wilk, L. Keoki Williams, Xiao Xu, Yifan Yang, Laura M. Yerges-Armstrong, Bing Yu, Faiez Zannad, Jing Hua Zhao, Sean L. Zheng, Simon de Denus, Antonio de Marvao, David van Heel, Jessica van Setten, Marion van Vugt, Pim van der Harst

#### Genes & Health Research Team

List of contributors from Genes & Health Research Team are available on: https://www.genesandhealth.org/research/scientific-publications-authorship-and-acknowledgments

At the time of the analysis, these include the following names (in alphabetical order by surname):

Shaheen Akhtar, Mohammad Anwar, Elena Arciero, Omar Asgar, Samina Ashraf, Saeed Bidi, Gerome Breen, James Broster, Raymond Chung, David Collier, Charles J Curtis, Shabana Chaudhary, Megan Clinch, Grainne Colligan, Panos Deloukas, Ceri Durham, Faiza Durrani, Fabiola Eto, Sarah Finer, Joseph Gafton, Ana Angel, Chris Griffiths, Joanne Harvey, Teng Heng, Sam Hodgson, Qin Qin Huang, Matt Hurles, Karen A Hunt, Shapna Hussain, Kamrul Islam, Vivek Iyer, Ben Jacobs, Ahsan Khan, Claudia Langenberg, Cath Lavery, Sang Hyuck Lee, Robin Lerner, Daniel MacArthur, Sidra Malik, Daniel Malawsky, Hilary Martin, Dan Mason, Rohini Mathur, Mohammed Bodrul Mazid, John McDermott, Caroline Morton, Bill Newman, Elizabeth Owor, Asma Qureshi, Samiha Rahman, Shwetha Ramachandrappa, Mehru Raza, Jessry Russell, Nishat Safa, Miriam Samuel, Michael Simpson, John Solly, Marie Spreckley. Daniel Stow, Michael Taylor, Richard C Trembath, Karen Tricker, Nasir Uddin, David A van Heel, Klaudia Walter, Caroline Winckley, Suzanne Wood, John Wright, Ishevanhu Zengeya, Julia Zöllner

#### DBDS Genomic Consortium

The following names are members of the Danish Blood Donor Study (DBDS) Genomic Consortium at the time of the analysis, listed in alphabetical order by surname:

Karina Banasik[1], Jakob Bay[2], Jens Kjærgaard Boldsen[3], Thorsten Brodersen[2], Søren Brunak[1], Kristoffer Burgdorf[1], Mona Ameri Chalmer[4], Maria Didriksen[5], Khoa Manh Dinh[3], Joseph Dowsett[5], Christian Erikstrup[3,6], Bjarke Feenstra[5,7], Frank Geller[5,7], Daniel Gudbjartsson[8], Thomas Folkmann Hansen[4], Lotte Hindhede[3], Henrik Hjalgrim[9,7], Rikke Louise Jacobsen[5], Gregor Jemec[10], Bitten Aagaard Jensen[11], Katrine Kaspersen[3], Bertram Dalskov Kjerulff[3], Lisette Kogelman[4], Margit Anita Hørup Larsen[5], Ioannis Louloudis[1], Agnete Lundgaard[1], Susan Mikkelsen[3], Christina Mikkelsen[5], Ioanna Nissen[5], Mette Nyegaard[12], Sisse Rye Ostrowski[5,13], Ole Birger Pedersen[2,13], Alexander Pil Henriksen[1], Palle Duun Rohde[12], Klaus Rostgaard[9,7], Michael Schwinn[5], Kari Stefansson[8], Hreinn Stefánsson[8], Erik Sørensen[5], Unnur Þorsteinsdóttir[8], Lise Wegner Thørner[5], Mie Topholm Bruun[14], Henrik Ullum[15], Thomas Werge[16,13], David Westergaard[1]

##### Affiliation

1. Novo Nordisk Foundation Center for Protein Research, Faculty of Health and Medical Sciences, University of Copenhagen, Copenhagen, Denmark
2. Department of Clinical Immunology, Zealand University Hospital, Køge, Denmark
3. Department of Clinical Immunology, Aarhus University Hospital, Aarhus, Denmark
4. Danish Headache Center, Department of Neurology, Copenhagen University Hospital, Rigshospitalet-Glostrup, Copenhagen, Denmark
5. Department of Clinical Immunology, Copenhagen University Hospital, Rigshospitalet, Copenhagen, Denmark
6. Department of Clinical Medicine, Health, Aarhus University, Aarhus, Denmark
7. Department of Epidemiology Research, Statens Serum Institut, Copenhagen, Denmark
8. deCODE Genetics, Reykjavik, Iceland
9. Danish Cancer Society Research Center, Copenhagen, Denmark
10. Department of Dermatology, Zealand University hospital, Roskilde, Denmark
11. Department of Clinical Immunology, Aalborg University Hospital, Aalborg, Denmark
12. Department of Health Science and Technology, Faculty of Medicine, Aalborg Univeristy, Aalborg, Denmark
13. Department of Clinical Medicine, Faculty of Health and Medical Sciences, University of Copenhagen, Copenhagen, Denmark
14. Department of Clinical Immunology, Odense University Hospital, Odense, Denmark
15. Statens Serum Institut, Copenhagen, Denmark
16. Institute of Biological Psychiatry, Mental Health Centre, Sct. Hans, Copenhagen University Hospital, Roskilde, Denmark

### APPENDICES

#### Appendix 1

Heart failure code list

| Source | Code | Description |
|---|---:|---|
| ICD10 | I50.0 | Congestive heart failure |
| ICD10 | I50.1 | Left ventricular failure |
| ICD10 | I50.2 | Systolic (congestive) heart failure |
| ICD10 | I50.4 | Combined systolic (congestive) and diastolic (congestive) heart failure |
| ICD10 | I50.9 | Heart failure, unspecified |
| ICD10 | I11.0 | Hypertensive heart disease with (congestive) heart failure |
| ICD10 | I13.0 | Hypertensive heart and renal disease with (congestive) heart failure |
| ICD10 | I13.2 | Hypertensive heart and renal disease with both (congestive) heart failure and renal failure |
| ICD9 | 428.* | Heart failure |
| ICD9 | 402.01 | Malignant hypertensive heart disease with heart failure |
| ICD9 | 402.11 | Benign hypertensive heart disease with heart failure |
| ICD9 | 402.91 | Unspecified hypertensive heart disease with heart failure |

**NOTE:**

- Code *XXX.** should include all child terms starting with *XXX*, e.g., *428.** should include 428.0, 428.1, 428.20, etc.
- Some code dictionaries do not have "." separator, so please adjust accordingly

**Appendix 2**

Heart failure medication list

| |
|---|
| furosemide |
| lasix |
| bumetanide |
| bumex |
| torsemide |
| demadex |
| ethacrynic acid |
| edecrin |
| metolazone |
| zaroxolyn |

**Appendix 3**

Free text search strings for heart failure

| Text Terms | Negation Terms |
|---|---|
| heart failure | family history |
| left ventricular failure | fhx |
| cardiomyopathy | mother |
| | mom |
| | father |
| | dad |
| | brother |
| | sister |
| | aunt |
| | uncle |
| | grandma |
| | grandpa |
| | no |
| | not |
| | negative |

**Appendix 4**

Code list for coronary artery disease, valvular or congenital heart disease

| Source | Code | Description | Phenotype class |
|---|---|---|---|
| ICD10 | I21.* | Acute myocardial infarction | CAD |
| ICD10 | I22.* | Subsequent myocardial infarction | CAD |
| ICD10 | I23.* | Certain current complications following acute myocardial infarction | CAD |
| ICD10 | I24.1 | Dressler's syndrome | CAD |
| ICD10 | I25.2 | Old myocardial infarction | CAD |
| ICD10 | I25.5 | Ischaemic cardiomyopathy | CAD |
| ICD10 | I25.6 | Silent myocardial ischaemia | CAD |
| ICD10 | 745.* | Bulbus cordis anomalies and anomalies of cardiac septal closure | Congential HD |
| ICD10 | Q20.* | Congenital malformations of cardiac chambers and connections | Congential HD |
| ICD10 | Q21.* | Congenital malformations of cardiac septa | Congential HD |
| ICD10 | Q22.* | Congenital malformations of pulmonary and tricuspid valves | Congential HD |
| ICD10 | Q23.* | Congenital malformations of aortic and mitral valves | Congential HD |
| ICD10 | Q24.* | Other congenital malformations of heart | Congential HD |
| ICD10 | Q25.* | Congenital malformations of great arteries | Congential HD |
| ICD10 | Q26.* | Congenital malformations of great veins | Congential HD |
| ICD10 | 746.* | Other congenital anomalies of heart | Valve disease |
| ICD10 | I05.* | Rheumatic mitral valve diseases | Valve disease |
| ICD10 | I06.* | Rheumatic aortic valve diseases | Valve disease |
| ICD10 | I07.* | Rheumatic tricuspid valve diseases | Valve disease |
| ICD9 | 410.* | Myocardial infarction | CAD |
| ICD9 | 412.* | Old myocardial infarction | CAD |
| ICD9 | 394.* | Diseases of mitral valve | Valve disease |
| ICD9 | 395.* | Diseases of aortic valve | Valve disease |
| ICD9 | 396.* | Diseases of mitral and aortic valves | Valve disease |
| ICD9 | 397.* | Diseases of other endocardial structures | Valve disease |
| OPCS4 | K40.* | Saphenous vein graft replacement of coronary artery | CAD |
| OPCS4 | K41.* | Other autograft replacement of coronary artery | CAD |
| OPCS4 | K42.* | Allograft replacement of coronary artery | CAD |
| OPCS4 | K43.* | Prosthetic replacement of coronary artery | CAD |
| OPCS4 | K44.* | Other replacement of coronary artery | CAD |
| OPCS4 | K45.* | Connection of thoracic artery to coronary artery | CAD |
| OPCS4 | K46.* | Other bypass of coronary artery | CAD |
| OPCS4 | K47.* | Repair of coronary artery | CAD |
| OPCS4 | K48.* | Other open operations on coronary artery | CAD |
| OPCS4 | K49.* | Transluminal balloon angioplasty of coronary artery | CAD |
| OPCS4 | K50.* | Other therapeutic transluminal operations on coronary artery | CAD |
| OPCS4 | K75.* | Percutaneous transluminal balloon angioplasty and insertion of stent into coronary artery | CAD |
| OPCS4 | K25.* | Plastic repair of mitral valve | Valve disease |
| OPCS4 | K26.* | Plastic repair of aortic valve | Valve disease |
| OPCS4 | K27.* | Plastic repair of tricuspid valve | Valve disease |
| OPCS4 | K28.* | Plastic repair of pulmonary valve | Valve disease |
| OPCS4 | K29.* | Plastic repair of unspecified valve of heart | Valve disease |

| | | | |
|---|---|---|---|
| OPCS4 | K30.* | Revision of plastic repair of valve of heart | Valve disease |
| OPCS4 | K31.* | Open incision of valve of heart | Valve disease |
| OPCS4 | K32.* | Closed incision of valve of heart | Valve disease |
| OPCS4 | K34.* | Other open operations on valve of heart | Valve disease |
| OPCS4 | K36.* | Excision of valve of heart | Valve disease |
| OPCS4 | K37.* | Removal of obstruction from structure adjacent to valve of heart | Valve disease |
| OPCS4 | K38.* | Other operations on structure adjacent to valve of heart | Valve disease |
| CPT4 | 33510 | Coronary artery bypass, vein only; single coronary venous graft | CAD |
| CPT4 | 33510 | Coronary artery bypass, vein only; single coronary venous graft | CAD |
| CPT4 | 33511 | Coronary artery bypass, vein only; 2 coronary venous grafts | CAD |
| CPT4 | 33511 | Coronary artery bypass, vein only; 2 coronary venous grafts | CAD |
| CPT4 | 33512 | Coronary artery bypass, vein only; 3 coronary venous grafts | CAD |
| CPT4 | 33512 | Coronary artery bypass, vein only; 3 coronary venous grafts | CAD |
| CPT4 | 33513 | Coronary artery bypass, vein only; 4 coronary venous grafts | CAD |
| CPT4 | 33513 | Coronary artery bypass, vein only; 4 coronary venous grafts | CAD |
| CPT4 | 33530 | Reoperation, coronary artery bypass procedure or valve procedure, more than 1 month after original operation (List separately in addition to code for primary procedure) | CAD |
| CPT4 | 33533 | Coronary artery bypass, using arterial graft(s); single arterial graft | CAD |
| CPT4 | 33533 | Coronary artery bypass, using arterial graft(s); single arterial graft | CAD |
| CPT4 | 33534 | Coronary artery bypass, using arterial graft(s); 2 coronary arterial grafts | CAD |
| CPT4 | 33534 | Coronary artery bypass, using arterial graft(s); 2 coronary arterial grafts | CAD |
| CPT4 | 33535 | Coronary artery bypass, using arterial graft(s); 3 coronary arterial grafts | CAD |
| CPT4 | 33535 | Coronary artery bypass, using arterial graft(s); 3 coronary arterial grafts | CAD |
| CPT4 | 33536 | Coronary artery bypass, using arterial graft(s); 4 or more coronary arterial grafts | CAD |
| CPT4 | 33536 | Coronary artery bypass, using arterial graft(s); 4 or more coronary arterial grafts | CAD |
| CPT4 | 92920 | Percutaneous transluminal coronary angioplasty; single major coronary artery or branch | CAD |
| CPT4 | 92920 | Percutaneous transluminal coronary angioplasty; single major coronary artery or branch | CAD |
| CPT4 | 92921 | Percutaneous transluminal coronary angioplasty; each additional branch of a major coronary artery | CAD |
| CPT4 | 92921 | Percutaneous transluminal coronary angioplasty; each additional branch of a major coronary artery | CAD |
| CPT4 | 92924 | Percutaneous transluminal coronary atherectomy, with coronary angioplasty when performed; single major coronary artery or branch | CAD |
| CPT4 | 92924 | Percutaneous transluminal coronary atherectomy, with coronary angioplasty when performed; single major coronary artery or branch | CAD |
| CPT4 | 92925 | Percutaneous transluminal coronary atherectomy, with coronary angioplasty when performed; each additional branch of a major coronary artery | CAD |
| CPT4 | 92925 | Percutaneous transluminal coronary atherectomy, with coronary angioplasty when performed; each additional branch of a major coronary artery | CAD |
| CPT4 | 92928 | Percutaneous transcatheter placement of intracoronary stent(s), with coronary angioplasty when performed; single major coronary artery or branch | CAD |
| CPT4 | 92928 | Percutaneous transcatheter placement of intracoronary stent(s), with coronary angioplasty when performed; single major coronary artery or branch | CAD |
| CPT4 | 92929 | Percutaneous transcatheter placement of intracoronary stent(s), with coronary angioplasty when performed; each additional branch of a major coronary artery | CAD |
| CPT4 | 92929 | Percutaneous transcatheter placement of intracoronary stent(s), with coronary angioplasty when performed; each additional branch of a major coronary artery | CAD |
| CPT4 | 92933 | Percutaneous transluminal coronary atherectomy, with intracoronary stent, with coronary angioplasty when performed; single major coronary artery or branch | CAD |
| CPT4 | 92933 | Percutaneous transluminal coronary atherectomy, with intracoronary stent, with coronary angioplasty when performed; single major coronary artery or branch | CAD |

| | | | |
|---|---|---|---|
| CPT4 | 92934 | Percutaneous transluminal coronary atherectomy, with intracoronary stent, with coronary angioplasty when performed; each additional branch of a major coronary artery | CAD |
| CPT4 | 92934 | Percutaneous transluminal coronary atherectomy, with intracoronary stent, with coronary angioplasty when performed; each additional branch of a major coronary artery | CAD |
| CPT4 | 92937 | Percutaneous transluminal revascularization of or through coronary artery bypass graft (internal mammary, free arterial, venous), any combination of intracoronary stent, atherectomy and angioplasty, including distal protection when performed; single vessel | CAD |
| CPT4 | 92937 | Percutaneous transluminal revascularization of or through coronary artery bypass graft (internal mammary, free arterial, venous), any combination of intracoronary stent, atherectomy and angioplasty, including distal protection when performed; single vessel | CAD |
| CPT4 | 92938 | Percutaneous transluminal revascularization of or through coronary artery bypass graft (internal mammary, free arterial, venous), any combination of intracoronary stent, atherectomy and angioplasty, including distal protection when performed; each additional branch subtended by the bypass graft | CAD |
| CPT4 | 92938 | Percutaneous transluminal revascularization of or through coronary artery bypass graft (internal mammary, free arterial, venous), any combination of intracoronary stent, atherectomy and angioplasty, including distal protection when performed; each additional branch subtended by the bypass graft | CAD |
| CPT4 | 92941 | Percutaneous transluminal revascularization of acute total/subtotal occlusion during acute myocardial infarction, coronary artery or coronary artery bypass graft, any combination of intracoronary stent, atherectomy and angioplasty, including aspiration thrombectomy when performed, single vessel | CAD |
| CPT4 | 92941 | Percutaneous transluminal revascularization of acute total/subtotal occlusion during acute myocardial infarction, coronary artery or coronary artery bypass graft, any combination of intracoronary stent, atherectomy and angioplasty, including aspiration thrombectomy when performed, single vessel | CAD |
| CPT4 | 92943 | Percutaneous transluminal revascularization of chronic total occlusion, coronary artery, coronary artery branch, or coronary artery bypass graft, any combination of intracoronary stent, atherectomy and angioplasty; single vessel | CAD |
| CPT4 | 92943 | Percutaneous transluminal revascularization of chronic total occlusion, coronary artery, coronary artery branch, or coronary artery bypass graft, any combination of intracoronary stent, atherectomy and angioplasty; single vessel | CAD |
| CPT4 | 92944 | Percutaneous transluminal revascularization of chronic total occlusion, coronary artery, coronary artery branch, or coronary artery bypass graft, any combination of intracoronary stent, atherectomy and angioplasty; each additional coronary artery, coronary artery branch, or bypass graft | CAD |
| CPT4 | 92944 | Percutaneous transluminal revascularization of chronic total occlusion, coronary artery, coronary artery branch, or coronary artery bypass graft, any combination of intracoronary stent, atherectomy and angioplasty; each additional coronary artery, coronary artery branch, or bypass graft | CAD |
| CPT4 | 92973 | Percutaneous transluminal coronary thrombectomy, mechanical* | CAD |
| CPT4 | 92973 | Percutaneous transluminal coronary thrombectomy, mechanical* | CAD |
| CPT4 | 92975 | Thrombolysis, coronary; by intracoronary infusion, including selective coronary angiography | CAD |
| CPT4 | 92975 | Thrombolysis, coronary; by intracoronary infusion, including selective coronary angiography | CAD |
| CPT4 | 33608 | Repair of complex cardiac anomaly other than pulmonary atresia with ventricular septal defect by construction or replacement of conduit from right or left ventricle to pulmonary artery | Congenital HD |
| CPT4 | 33610 | Repair of complex cardiac anomalies (eg, single ventricle with subaortic obstruction) by surgical enlargement of ventricular septal defect | Congenital HD |
| CPT4 | 33611 | Repair of double outlet right ventricle with intraventricular tunnel repair; | Congenital HD |
| CPT4 | 33612 | Repair of double outlet right ventricle with intraventricular tunnel repair; with repair of right ventricular outflow tract obstruction | Congenital HD |
| CPT4 | 33615 | Repair of complex cardiac anomalies (eg, tricuspid atresia) by closure of atrial septal defect and anastomosis of atria or vena cava to pulmonary artery (simple Fontan procedure) | Congenital HD |
| CPT4 | 33617 | Repair of complex cardiac anomalies (e.g., single ventricle by modified Fontan) | Congenital HD |

| | | | |
|---|---|---|---|
| CPT4 | 33619 | Repair of single ventricle with aortic outflow obstruction and aortic arch hypoplasia (hypoplastic left heart syndrome) (eg, Norwood procedure) | Congenital HD |
| CPT4 | 33641 | Repair atrial septal defect, secundum, with cardiopulmonary bypass, with or without patch | Congenital HD |
| CPT4 | 33645 | Direct or patch closure, sinus venosus, with or without anomalous pulmonary venous drainage | Congenital HD |
| CPT4 | 33647 | Repair of atrial septal defect and ventricular septal defect, with direct or patch closure | Congenital HD |
| CPT4 | 33660 | Repair of incomplete or partial atrioventricular canal (ostium primum atrial septal defect), with or without atrioventricular valve repair | Congenital HD |
| CPT4 | 33665 | Repair of intermediate or transitional atrioventricular canal, with or without atrioventricular valve repair | Congenital HD |
| CPT4 | 33670 | Repair of complete atrioventricular canal, with or without prosthetic valve | Congenital HD |
| CPT4 | 33675 | Closure of multiple ventricular septal defects; | Congenital HD |
| CPT4 | 33676 | Closure of multiple ventricular septal defects; with pulmonary valvotomy or infundibular resection (acyanotic) | Congenital HD |
| CPT4 | 33677 | Closure of multiple ventricular septal defects; with removal of pulmonary artery band, with or without gusset | Congenital HD |
| CPT4 | 33681 | Closure of single ventricular septal defect, with or without patch; | Congenital HD |
| CPT4 | 33684 | Closure of single ventricular septal defect, with or without patch; with pulmonary valvotomy or infundibular resection (acyanotic) | Congenital HD |
| CPT4 | 33688 | Closure of single ventricular septal defect, with or without patch; with removal of pulmonary artery band, with or without gusset | Congenital HD |
| CPT4 | 33692 | Complete repair tetralogy of Fallot without pulmonary atresia; | Congenital HD |
| CPT4 | 33694 | Complete repair tetralogy of Fallot without pulmonary atresia; with transannular patch | Congenital HD |
| CPT4 | 33697 | Complete repair tetralogy of Fallot with pulmonary atresia including construction of conduit from right ventricle to pulmonary artery and closure of ventricular septal defect | Congenital HD |
| CPT4 | 33702 | Repair sinus of Valsalva fistula, with cardiopulmonary bypass; | Congenital HD |
| CPT4 | 33710 | Repair sinus of Valsalva fistula, with cardiopulmonary bypass; with repair of ventricular septal defect | Congenital HD |
| CPT4 | 33720 | Repair sinus of Valsalva aneurysm, with cardiopulmonary bypass | Congenital HD |
| CPT4 | 33722 | Closure of aortico-left ventricular tunnel | Congenital HD |
| CPT4 | 33732 | Repair of cor triatriatum or supravalvular mitral ring by resection of left atrial membrane | Congenital HD |
| CPT4 | 33735 | Atrial septectomy or septostomy; closed heart (Blalock-Hanlon type operation) | Congenital HD |
| CPT4 | 33736 | Atrial septectomy or septostomy; open heart with cardiopulmonary bypass | Congenital HD |
| CPT4 | 33737 | Atrial septectomy or septostomy; open heart, with inflow occlusion | Congenital HD |
| CPT4 | 33770 | Repair of transposition of the great arteries with ventricular septal defect and subpulmonary stenosis; without surgical enlargement of ventricular septal defect | Congenital HD |
| CPT4 | 33774 | Repair of transposition of the great arteries, atrial baffle procedure (eg, Mustard or Senning type) with cardiopulmonary bypass; | Congenital HD |
| CPT4 | 33776 | Repair of transposition of the great arteries, atrial baffle procedure (eg, Mustard or Senning type) with cardiopulmonary bypass; with closure of ventricular septal defect | Congenital HD |
| CPT4 | 33780 | Repair of transposition of the great arteries, aortic pulmonary artery reconstruction (eg, Jatene type); with closure of ventricular septal defect | Congenital HD |
| CPT4 | 33782 | Aortic root translocation with ventricular septal defect and pulmonary stenosis repair (ie, Nikaidoh procedure); without coronary ostium reimplantation | Congenital HD |
| CPT4 | 33783 | Aortic root translocation with ventricular septal defect and pulmonary stenosis repair (ie, Nikaidoh procedure); with reimplantation of 1 or both coronary ostia | Congenital HD |
| CPT4 | 33786 | Total repair, truncus arteriosus (Rastelli type operation) | Congenital HD |
| CPT4 | 33813 | Obliteration of aortopulmonary septal defect; without cardiopulmonary bypass | Congenital HD |
| CPT4 | 33814 | Obliteration of aortopulmonary septal defect; with cardiopulmonary bypass | Congenital HD |
| CPT4 | 33920 | Repair of pulmonary atresia with ventricular septal defect, by construction or replacement of conduit from right or left ventricle to pulmonary artery | Congenital HD |
| CPT4 | 33361 | Transcatheter aortic valve replacement (TAVR/TAVI) with prosthetic valve; percutaneous femoral artery approach | Valve disease |

| | | | |
|---|---|---|---|
| CPT4 | 33362 | Transcatheter aortic valve replacement (TAVR/TAVI) with prosthetic valve; open femoral artery approach | Valve disease |
| CPT4 | 33363 | Transcatheter aortic valve replacement (TAVR/TAVI) with prosthetic valve; open axillary artery approach | Valve disease |
| CPT4 | 33364 | Transcatheter aortic valve replacement (TAVR/TAVI) with prosthetic valve; open iliac artery approach | Valve disease |
| CPT4 | 33365 | Transcatheter aortic valve replacement (TAVR/TAVI) with prosthetic valve; transaortic approach (e.g., median sternotomy, mediastinotomy) | Valve disease |
| CPT4 | 33366 | Transcatheter aortic valve replacement (TAVR/TAVI) with prosthetic valve; transapical exposure (e.g., left thoracotomy) | Valve disease |
| CPT4 | 33367 | Transcatheter aortic valve replacement (TAVR/TAVI) with prosthetic valve; cardiopulmonary bypass support with percutaneous peripheral arterial and venous cannulation (e.g., femoral vessels) (List separately in addition to code for primary procedure) | Valve disease |
| CPT4 | 33368 | Transcatheter aortic valve replacement (TAVR/TAVI) with prosthetic valve; cardiopulmonary bypass support with open peripheral arterial and venous cannulation (e.g., femoral, iliac, axillary vessels) (List separately in addition to code for primary procedure) | Valve disease |
| CPT4 | 33369 | Transcatheter aortic valve replacement (TAVR/TAVI) with prosthetic valve; cardiopulmonary bypass support with central arterial and venous cannulation (e.g., aorta, right atrium, pulmonary artery) (List separately in addition to code for primary procedure) | Valve disease |
| CPT4 | 33390 | Valvuloplasty, aortic valve, open, with cardiopulmonary bypass; simple (ie, valvotomy, debridement, debulking and/or simple commissural resuspension) | Valve disease |
| CPT4 | 33391 | Valvuloplasty, aortic valve, open, with cardiopulmonary bypass; complex (eg, leaflet extension, leaflet resection, leaflet reconstruction or annuloplasty) | Valve disease |
| CPT4 | 33400 | Aortic valvuloplasty | Valve disease |
| CPT4 | 33401 | Open valvuloplasty of aortic valve with inflow occlusion | Valve disease |
| CPT4 | 33403 | Valvuloplasty, aortic valve; using transventricular dilation, with cardiopulmonary bypass | Valve disease |
| CPT4 | 33404 | LV—aorta conduit | Valve disease |
| CPT4 | 33405 | Replacement, aortic valve, with cardiopulmonary bypass; with prosthetic valve other than homograft or stentless valve | Valve disease |
| CPT4 | 33406 | Replacement, aortic valve, with cardiopulmonary bypass; with allograft valve (freehand) | Valve disease |
| CPT4 | 33410 | Replacement, aortic valve, with cardiopulmonary bypass; with stentless tissue valve | Valve disease |
| CPT4 | 33411 | Replacement, aortic valve; with aortic annulus enlargement, noncoronary sinus | Valve disease |
| CPT4 | 33412 | Replacement, aortic valve; with transventricular aortic annulus enlargement (Konno procedure) | Valve disease |
| CPT4 | 33413 | Replacement, aortic valve; by translocation of autologous pulmonary valve with allograft replacement of pulmonary valve (Ross procedure) | Valve disease |
| CPT4 | 33415 | Resection or incision of subvalvular tissue for discrete subvalvular aortic stenosis | Valve disease |
| CPT4 | 33418 | Transcatheter mitral valve repair, percutaneous approach, including transseptal | Valve disease |
| CPT4 | 33419 | Transcatheter mitral valve repair, percutaneous approach, including transseptal puncture when performed; additional prosthesis(es) during same session (List separately in addition to code for primary procedure) | Valve disease |
| CPT4 | 33420 | Valvotomy, mitral valve; closed heart | Valve disease |
| CPT4 | 33422 | Valvotomy, mitral valve; open heart, with cardiopulmonary bypass | Valve disease |
| CPT4 | 33425 | Valvuloplasty, mitral valve, with cardiopulmonary bypass | Valve disease |
| CPT4 | 33426 | Valvuloplasty, mitral valve, with cardiopulmonary bypass; with prosthetic ring | Valve disease |
| CPT4 | 33427 | Valvuloplasty, mitral valve, with cardiopulmonary bypass; radical reconstruction, with or without ring | Valve disease |
| CPT4 | 33430 | Replacement, mitral valve, with cardiopulmonary bypass | Valve disease |
| CPT4 | 33440 | Replacement of aortic valve by translocation of autologous pulmonary valve and transventricular aortic annulus enlargement of left ventricular outflow tract with valved conduit replacement of pulmonary valve | Valve disease |
| CPT4 | 33460 | Valvectomy, tricuspid valve, with cardiopulmonary bypass | Valve disease |

| | | | |
|---|---|---|---|
| CPT4 | 33463 | Valvuloplasty, tricuspid valve; without ring insertion | Valve disease |
| CPT4 | 33464 | Valvuloplasty, tricuspid valve; with ring insertion | Valve disease |
| CPT4 | 33465 | Replacement, tricuspid valve, with cardiopulmonary bypass | Valve disease |
| CPT4 | 33468 | Tricuspid valve repositioning and plication for Ebstein anomaly | Valve disease |
| CPT4 | 33470 | Valvotomy, pulmonary valve, closed heart; transventricular | Valve disease |
| CPT4 | 33471 | Valvotomy, pulmonary valve, closed heart; via pulmonary artery | Valve disease |
| CPT4 | 33472 | Incision of valve at right lower heart chamber, open procedure | Valve disease |
| CPT4 | 33474 | Valvotomy, pulmonary valve, open heart, with cardiopulmonary bypass | Valve disease |
| CPT4 | 33475 | Replacement, pulmonary valve | Valve disease |
| CPT4 | 33476 | Right ventricular resection for infundibular stenosis, with or without commissurotomy | Valve disease |
| CPT4 | 33477 | Transcatheter pulmonary valve implantation, percutaneous approach, including prestenting of the valve delivery site, when performed | Valve disease |
| CPT4 | 33496 | Repair of non-structural prosthetic valve dysfunction with cardiopulmonary bypass (separate procedure) | Valve disease |
| CPT4 | 33600 | Closure of atrioventricular valve (mitral or tricuspid) by suture or patch | Valve disease |
| CPT4 | 33602 | Closure of semilunar valve (aortic or pulmonary) by suture or patch | Valve disease |
| CPT4 | 92986 | Percutaneous balloon valvuloplasty; aortic valve | Valve disease |
| CPT4 | 92987 | Percutaneous balloon valvuloplasty; mitral valve | Valve disease |
| CPT4 | 92990 | Percutaneous balloon valvuloplasty; pulmonary valve | Valve disease |
| CPT4 | 93355 | Echocardiography, transesophageal (TEE) for guidance of a transcatheter intracardiac or great vessel(s) structural intervention(s) (eg, TAVR, transcatheter pulmonary valve replacement, mitral valve repair, paravalvular regurgitation repair, left atrial ap | Valve disease |
| CPT4 | 93590 | Percutaneous transcatheter closure of paravalvular leak; initial occlusion device, mitral valve | Valve disease |
| CPT4 | 93592 | Percutaneous transcatheter closure of paravalvular leak; each additional occlusion device (List separately in addition to code for primary procedure) | Valve disease |
| CPT4 | 0256T | Implantation of catheter-delivered prosthetic aortic heart valve; endovascular approach | Valve disease |
| CPT4 | 0257T | Implantation of catheter-delivered prosthetic aortic heart valve; open thoracic approach (eg, transapical, transventricular) | Valve disease |
| CPT4 | 0262T | Implantation of catheter-delivered prosthetic pulmonary valve, endovascular approach | Valve disease |
| CPT4 | 0318T | Implantation of catheter-delivered prosthetic aortic heart valve, open thoracic approach, (eg, transapical, other than transaortic) | Valve disease |
| CPT4 | 0343T | Transcatheter mitral valve repair percutaneous approach including transseptal puncture when performed; initial prosthesis | Valve disease |
| CPT4 | 0345T | Transcatheter mitral valve repair percutaneous approach via the coronary sinus | Valve disease |
| CPT4 | 0483T | Transcatheter mitral valve implantation/replacement (TMVI) with prosthetic valve; percutaneous approach, including transseptal puncture, when performed | Valve disease |
| CPT4 | 0484T | Transcatheter mitral valve implantation/replacement (TMVI) with prosthetic valve; transthoracic exposure (e.g., thoracotomy, transapical) | Valve disease |

**NOTE:**

- Code *XXX.** should include all child terms starting with *XXX*, e.g., *Q20.** should include Q20.0, Q20.1, Q20.2, etc.
- Some code dictionaries do not have "." separator, so please adjust accordingly

**Appendix 5**

Code list for left ventricular systolic dysfunction (LVSD)

| Source | Code | Description |
|---|---:|---|
| ICD10 | I42.0 | Dilated cardiomyopathy (Congestive cardiomyopathy) |
| ICD10 | I42.6 | Alcoholic cardiomyopathy |
| ICD10 | I42.7 | Cardiomyopathy due to drug and external agent |
| ICD10 | I25.5 | Ischaemic cardiomyopathy |
| ICD10 | I50.2 | Systolic (congestive) heart failure |
| ICD10 | O90.3 | Cardiomyopathy in the puerperium |
| ICD9 | 428.2 | Systolic heart failure |
| ICD9 | 674.5 | Peripartum cardiomyopathy |
| ICD9 | 425.5 | Alcoholic cardiomyopathy |
| ICD9 | 425.9 | Secondary cardiomyopathy, unspecified |
| OPCS4 | K61.7 | Cardiac resynchronisation device |
| CPT4 | 33224 | Insertion of pacing electrode, cardiac venous system, for left ventricular pacing, with attachment to previously placed pacemaker or implantable defibrillator pulse generator (including revision of pocket, removal, insertion, and/or replacement of existing generator) |
| CPT4 | 33225 | Insertion of pacing electrode, cardiac venous system, for left ventricular pacing, at time of insertion of implantable defibrillator or pacemaker pulse generator (e.g., for upgrade to dual chamber system) (List separately in addition to code for primary procedure) |
| CPT4 | 33226 | Repositioning of previously implanted cardiac venous system (left ventricular) electrode (including removal, insertion and/or replacement of existing generator) |

**Appendix 6**

**Table 2.1.** Boolean algebra to define HF phenotypes. Phenotype classifiers are combined with a conjunction (i.e. AND) operator. For example, a participant with HF = TRUE AND CAD, valve, or cong HD = FALSE AND Ever LVSD = TRUE AND LVEF ≥50% = TRUE would be categorised as a Case for Phenotype 1, 2, and 3, but Excluded from Phenotype 4 analysis.

| Phenotype classifier | | | | Category |
|---|---|---|---|---|
| **HF** | **Secondary HF** | **Ever LVSD** | **LVEF ≥50%** | |
| **Phenotype 1: Heart failure** | | | | |
| TRUE | ANY | ANY | ANY | Case |
| FALSE | ANY | ANY | ANY | Control |
| **Phenotype 2: Non-ischaemic HF** | | | | |
| ANY | TRUE | ANY | ANY | Exclude |
| TRUE | FALSE | ANY | ANY | Case |
| FALSE | FALSE | ANY | ANY | Control |
| **Phenotype 3: Non-ischaemic HFrEF (LVEF <50%)** | | | | |
| ANY | TRUE | ANY | ANY | Exclude |
| TRUE | FALSE | FALSE | ANY | Exclude |
| TRUE | FALSE | TRUE | ANY | Case |
| FALSE | FALSE | ANY | ANY | Control |
| **Phenotype 4: Non-ischaemic HFpEF (LVEF ≥50%)** | | | | |
| ANY | TRUE | ANY | ANY | Exclude |
| TRUE | FALSE | FALSE | FALSE | Exclude |
| TRUE | FALSE | FALSE | TRUE | Case |
| FALSE | FALSE | ANY | ANY | Control |

#### Appendix 7

Schematic diagram of HF phenotyping algorithm

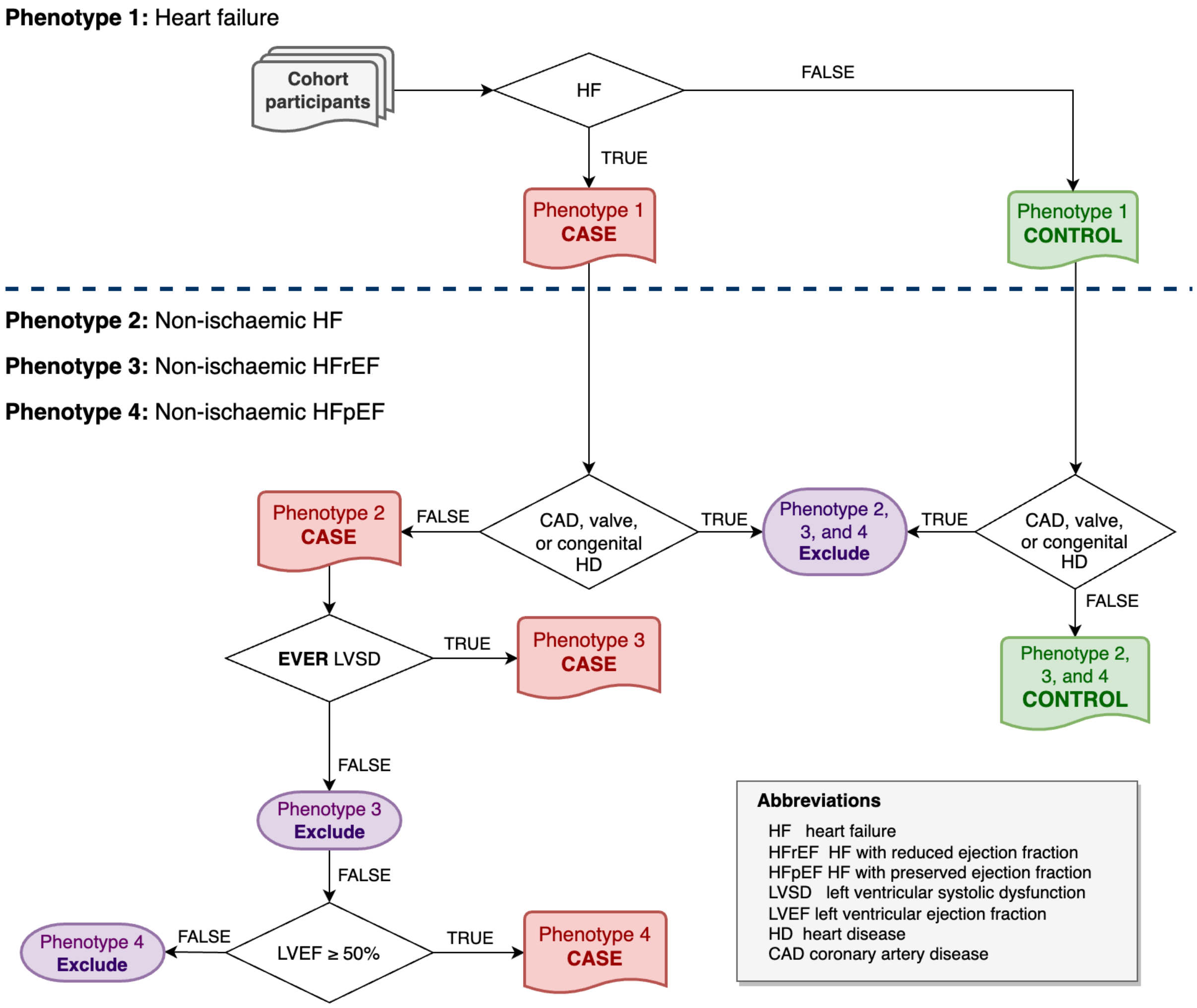
